# Genomic epidemiology as a tool for understanding drivers of hepatitis A community outbreaks in Massachusetts and New Hampshire

**DOI:** 10.64898/2026.05.14.26352933

**Authors:** Lydia A Krasilnikova, Lindsay Bouton, Taylor M Brock-Fisher, Emily Decker, Mary Godec, Zachary Thompson, Emily Dart, Fengxiang Gao, Adrianne Gladden-Young, Katelyn S Messer, Joshua Norville, Ivan Specht, Anthony Osinski, Jinfeng Li, Carrie Lones, Katherine C DeRuff, Katherine J Siddle, Daniel Church, Christopher Benton, Katrina Hansen, Hannah Bowen, Sanjib Bhattacharyya, Nicolas Epie, Catherine M Brown, Lawrence C Madoff, Bronwyn L MacInnis, Glen R Gallagher, Sandra Smole, Christine Bean, Elizabeth A Talbot, Meagan Burns, Matthew Doucette, Esther Fortes, Daniel J Park, Pardis C Sabeti, Shirlee Wohl

**Affiliations:** Massachusetts Department of Public Health, Boston, MA, USA; Broad Institute of MIT and Harvard, Cambridge, MA, USA; Department of Organismic and Evolutionary Biology, Harvard University, Cambridge, MA, USA; New Hampshire Division of Public Health, Department of Health and Human Services, Concord, NH, USA; Department of Molecular Microbiology and Immunology, Brown University, Providence, RI, USA; Howard Hughes Medical Institute, Chevy Chase, MD, USA; Department of Immunology and Infectious Disease, Harvard TH Chan School of Public Health, Boston, MA, USA; Division of Infectious Diseases, Brigham and Women’s Hospital, Harvard Medical School, Boston, MA, USA

## Abstract

Despite the existence of an effective vaccine, the United States continues to experience outbreaks of hepatitis A, including in Massachusetts (MA) and New Hampshire (NH) in 2018 and again in MA in 2023. To clarify the relationship between these outbreaks and better understand their drivers, we generated hepatitis A virus whole genome sequences from reported cases and analyzed them using open-source genotyping tools developed and released as part of this study. We found that the 2018 and 2023 outbreaks were caused by distinct viral strains, despite affecting individuals with similar demographic characteristics and reported risk factors. Detailed analysis of genomic and epidemiologic data further resolved transmission patterns within and across outbreaks, showing that experiencing homelessness and prior use of drugs were associated with increased transmission while also revealing transmission between individuals with and without these risk factors, as well as spread across state borders. Together, these findings demonstrate the value of broadly accessible genomic tools for understanding hepatitis A outbreaks and illustrate how whole genome analysis can complement epidemiological investigation by resolving transmission patterns and outbreak drivers that can inform public health interventions.

## Introduction

Hepatitis A, caused by the hepatitis A virus (HAV), remains widespread worldwide, with over 150 million infections annually ^1^. The virus is primarily spread via fecal-oral transmission and, while commonly asymptomatic in children, usually causes more severe disease in adults, with symptoms including fever, jaundice, nausea, vomiting, and diarrhea ^2^. Following the introduction of the hepatitis A vaccine in 1995 and its incorporation as a routine childhood vaccine throughout the United States (US) in 2006 ^2^, HAV infections declined by 97% from 1995 to 2015 ^3^. However, there remains a large population of adults who have neither been infected nor vaccinated; as of 2017, only 10.9% of US adults aged ≥19 had received hepatitis A vaccination ^2^. This susceptibility has contributed to continued HAV transmission among immunologically naive adults, culminating in a large outbreak beginning in 2016 that infected 44,930 individuals across 37 US states as of 2023 ^4^. This large outbreak was mostly associated with adults with known risk factors, particularly people who have used drugs (PWUD) and people experiencing homelessness (PEH). Specifically, across 33 states with available data, 53% of cases were in PWUD and 14% were in PEH. These risk factors are often associated with poor sanitation and overcrowded living conditions, which facilitate fecal-oral pathogen transmission. That said, this large outbreak stands out in the context of many previous HAV outbreaks, which are typically linked to contaminated foods rather than associated with specific demographic groups ^5^.

The Massachusetts Department of Public Health (MA DPH) confirmed 561 outbreak-associated hepatitis A cases (defined as cases not tied to foodborne spread or international travel) ^6,7^ between April 2018 and April 2020, many of which were associated with PWUD or PEH. The MA outbreak was characterized by unusually high morbidity and mortality, with a 79% hospitalization rate and 1.6% case fatality rate. This was likely due to underlying health conditions prevalent in the affected population, including a 46% hepatitis C coinfection rate. After 2020, hepatitis A case counts in MA returned to pre-outbreak levels, but there was a resurgence in 2023-2024, with 24 outbreak-associated cases reported using a narrower outbreak definition of only cases associated with experiencing homelessness, drug use, and recent incarceration (see **Methods** for detailed outbreak definitions). It was unclear if these cases were epidemiologically connected to the 2018–2020 outbreak. Furthermore, during the broader 2018-2024 period, there were an additional 169 hepatitis A cases in MA that did not meet these outbreak definitions, leaving uncertainty about the relationship between outbreak-associated cases and those in individuals without the same underlying risk factors. Our understanding of the outbreak is further complicated by the fact that the United States Centers for Disease Control and Prevention (US CDC) estimates that only half of HAV infections are typically reported to public health ^8^, and that under-reporting may be even more significant in PWUD and PEH^9^.

Vaccination is a cornerstone of HAV outbreak response and was heavily promoted during both the 2018-2020 ^10^ and 2023-2024 outbreaks in MA ^11^. Since 1996, adult hepatitis A vaccination recommendations in the US have been risk-based, targeting international travelers, men who have sex with men (MSM), PWUD, people with chronic liver disease, people living with HIV, and, beginning in 2019, PEH ^12^. Vaccination is also recommended as post-exposure prophylaxis (PEP) following known HAV exposure ^13^. Expanded access to hygiene (historically targeted to food handlers) can also mitigate HAV transmission, though such interventions can be resource-intensive ^14,15^. Understanding which populations are most affected and what drives HAV transmission is therefore critical for targeting vaccination efforts and evaluating the relative costs and benefits of intervention strategies.

In the US, HAV transmission is monitored in part by genotyping a subset of reported cases (typically <3%) using the US Centers for Disease Control and Prevention (CDC) Global Hepatitis Outbreak and Surveillance Technology (GHOST) system ^16^. Using this system involves sequencing a 349-nucleotide segment of the HAV genome called the VP1-2B region (also referred to as VP1-P2B or VP1-pX-2B) ^16,17^, then assigning a subgenotype-level identity ^18^ and a haplotype identifier using the GHOST internal typing system. The resulting profiles, as well as phylogenetic context, are used to assess relatedness between cases and contextualize outbreaks nationally. Whole genome sequencing, which may offer greater insight into transmission patterns, is not routinely performed.

In this context, key questions remain about the drivers and dynamics of recent HAV transmission. The US CDC Advisory Committee on Immunization Practices (ACIP) recognized drug use as a risk factor for hepatitis A in 1996 and experiencing homelessness in 2019 ^12,19^, but it remains unclear whether these risk factors are driving transmission. Relatedly, the degree to which HAV spreads across geographic boundaries and between populations with and without known risk factors remains poorly understood. In addition, it is unclear how hepatitis A vaccination practices in MA may have changed during and after the COVID-19 pandemic, and how such changes could shape future HAV transmission and outbreak risk.

To clarify HAV transmission patterns across time, geography, and risk groups that are not resolvable with routine subgenotyping, we attempted whole genome sequencing on all available MA HAV specimens and ultimately generated 226 HAV whole genome sequences from cases reported during 2018-2020 and 15 from 2023-2024. We also generated 61 HAV whole genome sequences from cases in neighboring New Hampshire (NH) from 2018-2020 to examine broader movement patterns and cross-border transmission. We paired MA and NH HAV genomic data with epidemiological information collected by MA DPH and the NH Division of Public Health (NH DPH), including demographic data, geographic data, reported risk factors, vaccination status, symptoms, travel history, and epidemiologic links. As part of this effort, we evaluated and developed methods for generating and characterizing HAV genomes, with the goal of providing broadly accessible tools to support improved investigation of future outbreaks.

## Results

### Epidemiological investigation of MA and NH outbreaks

We used detailed epidemiological data to explore risk factors among hepatitis A cases. In the 561 outbreak-associated cases in MA during 2018-2020, we observed high rates of PEH (N=181; 32.3%) and PWUD (N=303; 54.0%), with many reporting both risk factors (**Table 1**, **Fig S1**). We observed similar patterns in NH: among 338 confirmed HAV infections reported to the NH DPH between November 2018 and May 2020, 102 (30%) were in PEH and 185 (55%) were in PWUD. During the 2023-2024 resurgence in MA, when the definition of an outbreak-associated case was revised to include only cases with known risk factors (see **Methods**), 16 (66.7%) occurred among PEH and 18 (75.0%) among PWUD. Across the full 2018-2024 period, almost all MA cases occurred in individuals 18 years and older, consistent with the expected protective effect of childhood hepatitis A vaccination, which exceeded 90% coverage among MA adolescents by 2024. Only 2.2% of outbreak-associated cases in MA during this period occurred among known MSM, despite MSM having been a prominent risk factor in previous US hepatitis A outbreaks.

**Table 1.** Epidemiological characteristics of hepatitis A cases reported in MA and NH, 2018-2024. Outbreak-associated cases are those meeting state and CDC hepatitis A outbreak definitions. Successfully sequenced cases include all samples that generated high-quality HAV genome sequences. Not all outbreak-associated cases were successfully sequenced, and sequenced cases include both outbreak-associated and non-outbreak-associated cases.

|  | 2018-2020 |  |  | 2023-2024 |  | 2018-2024 |
|---|---|---|---|---|---|---|
|  | Outbreak-associated MA cases: % (N=X) | Successfully sequenced MA cases: % (N=X)<sup>1</sup> | Outbreak-associated NH cases: % (N=X) | Outbreak-associated MA cases: % (N=X) | Successfully sequenced MA cases: % (N=X)<sup>1</sup> | All MA cases: % (N=X)<sup>2</sup> |
| Total cases | N=561 | N=226 | N=338 (62 sequenced) | N=24 | N=15 | N=751 |
| Gender |  |  |  |  |  |  |
| Female | 36.2% (203) | 34.5% (78) | 42.6% (144) | 50% (12) | 53.3% (8) | 39.5% (297) |
| Male | 63.8% (358) | 65.5% (148) | 57.4% (194) | 50% (12) | 46.7% (7) | 60.5% (454) |
| Unknown | 0% (0) | 0% (0) | 0% (0) | 0% (0) | 0% (0) | 0% (0) |
| Age group (yrs) |  |  |  |  |  |  |
| <20 years old | 0.7% (4) | 0.9% (2) | 0.9% (3) | 0% (0) | 0% (0) | 1.7% (13) |
| 20-39 years old | 59.2% (332) | 61.9% (140) | 61.2% (207) | 37.5% (9) | 50% (4) | 54.2% (407) |
| 40-59 years old | 29.1% (163) | 28.8% (65) | 30.5% (103) | 58.3% (14) | 50% (4) | 28.6% (215) |
| >=60 years old | 11.1% (62) | 8.4% (19) | 7.1% (24) | 4.2% (1) | 0% (0) | 15.4% (116) |
| Unknown | 0% (0) | 0% (0) | 7.1% (24) | 0% (0) | 0% (0) | 0% (0) |
| Race/ethnicity combined |  |  |  |  |  |  |
| White | 67.2% (377) | 66.4% (150) | 83.1% (281) | 66.7% (16) | 46.7% (7) | 63.4% (476) |
| Hispanic/Latinx | 7.1% (40) | 9.3% (21) | 1.5% (5) | 16.7% (4) | 13.3% (2) | 8.5% (64) |
| Black/African American | 3.9% (22) | 5.8% (13) | 0.6% (2) | 8.3% (2) | 13.3% (2) | 4.9% (37) |
| Asian | 0.7% (4) | 0.4% (1) | 0% (0) | 0% (0) | 20% (3) | 2.9% (22) |
| Multirace | 5.9% (33) | 4.9% (11) | 1.2% (4) | 8.3% (2) | 0% (0) | 5.3% (40) |
| Other<sup>3</sup> | 2.9% (16) | 2.2% (5) | 4.1% (14) | 0% (0) | 0% (0) | 3.7% (28) |
| Unknown | 12.3% (69) | 11.1% (25) | 9.5% (32) | 0% (0) | 6.7% (1) | 11.2% (84) |
| History of drug use |  |  |  |  |  |  |
| Yes | 54% (303) | 53.5% (121) | 54.7% (185) | 75% (18) | 40% (6) | 44.9% (337) |
| No | 32.6% (183) | 32.7% (74) | 29% (98) | 8.3% (2) | 46.7% (7) | 40.2% (302) |
| Unknown | 13.4% (75) | 13.7% (31) | 16.3% (55) | 16.7% (4) | 13.3% (2) | 14.9% (112) |
| Experiencing homelessness |  |  |  |  |  |  |
| Yes | 32.3% (181) | 38.1% (86) | 30.2% (102) | 66.7% (16) | 33.3% (5) | 27.2% (204) |
| No | 56.1% (315) | 54% (122) | 51.5% (174) | 25% (6) | 60% (9) | 61.4% (461) |
| Unknown | 11.6% (65) | 8% (18) | 18.3% (62) | 8.3% (2) | 6.7% (1) | 11.5% (86) |
| Vaccinated |  |  |  |  |  |  |
| Yes | 4.1% (23) | 4% (9) | 8.3% (28) | 4.2% (1) | 0% (0) | 3.9% (29) |
| No/unknown | 95.9% (538) | 96% (217) | 91.7% (310) | 95.8% (23) | 100% (15) | 96.1% (722) |

<sup>1</sup>Successfully sequenced cases are not a subset of outbreak-associated cases; successfully sequenced cases contain both outbreak-associated and non-outbreak cases.
One sequenced case in 2018-2020 and 7 sequenced cases in 2023-2024 were not outbreak-associated.
<sup>2</sup>Contains all cases for the full 2018-2024 period, regardless of outbreak association or sequencing success.
<sup>3</sup>To avoid identifiability due to low numbers, individuals identifying as American Indian/Alaska Native and Native Hawaiian/Pacific Islander have been grouped with those that reported Other as race/ethnicity.

Due to the central role vaccination plays in hepatitis A outbreak response, we also explored adult vaccination patterns during the 2018-2020 and 2023-2024 outbreak periods. Very few MA cases in our dataset had ever been vaccinated (**Table 1**), so we focused this analysis on population-level vaccination trends rather than individual-level effects. Using data from the Massachusetts Immunization Information System (MIIS), we observed that MA hepatitis A case counts declined following targeted vaccination efforts initiated in 2018. However, overall adult hepatitis A vaccination rates decreased after the onset of the COVID-19 pandemic and did not return to pre-pandemic levels during the study period (**Fig 1A**). Although some decline in adult vaccination coverage is expected as cohorts vaccinated in childhood age into adulthood, this cohort effect alone is unlikely to explain the magnitude of the observed decline, particularly given similar trends among older age groups.

**Figure 1.**
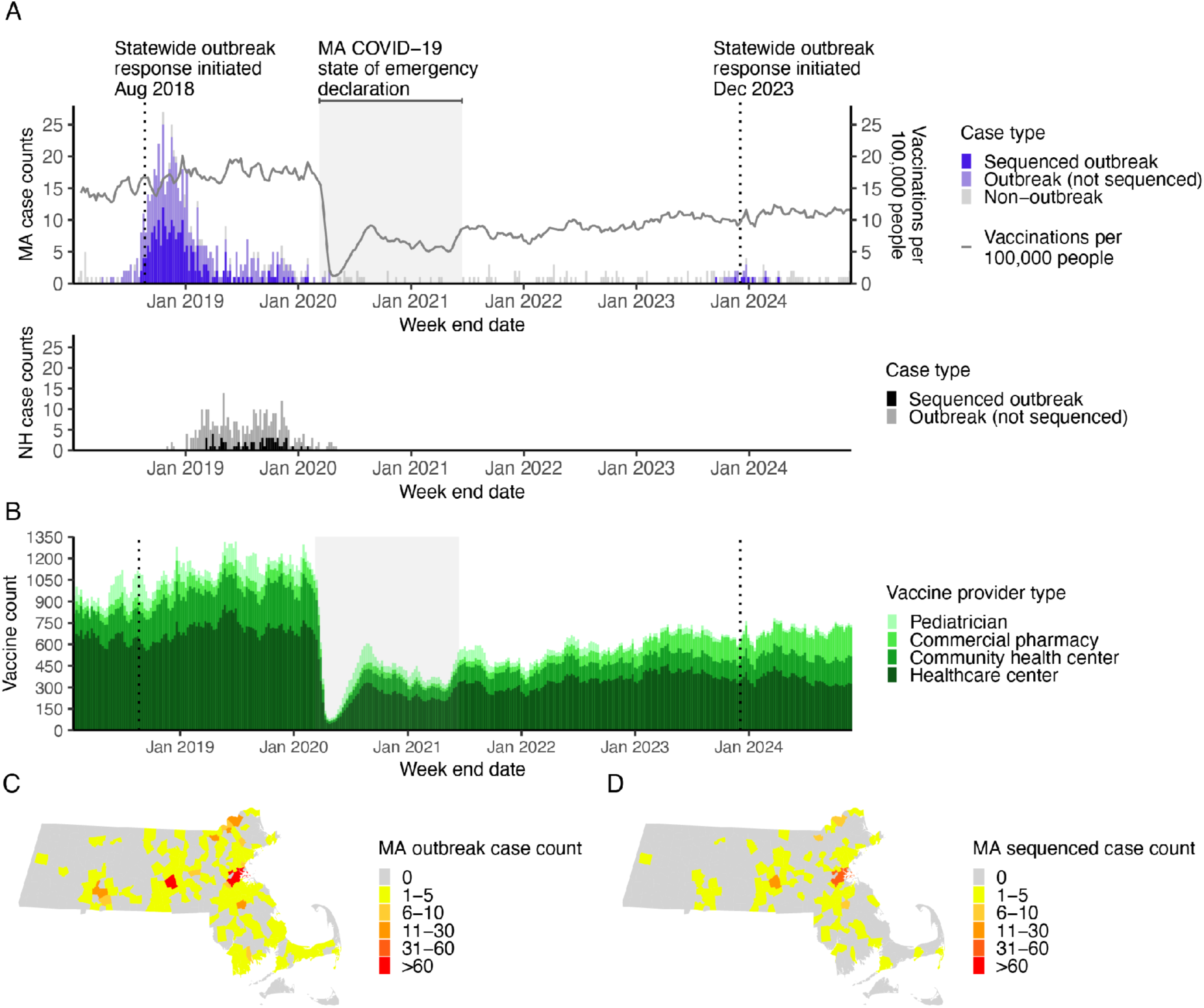
Total confirmed and sequenced HAV cases from late 2018 through early 2024. **A)** Top: Total confirmed cases in MA, shaded by whether cases were successfully sequenced in this study (left axis) and HAV vaccines administered in MA per 100,000 people (right axis). Bottom: outbreak-associated cases in NH, shaded by whether cases were successfully sequenced in this study. **B)** HAV vaccines administered by vaccine provider type. **C)** Total outbreak-associated cases by MA town. **D)** Total sequenced cases by MA town, including both outbreak-associated and non-outbreak-associated cases.

Furthermore, most of the adult population in our study period is too old to have benefited from universal childhood hepatitis A vaccine recommendations (established in 2006). Finally, populations at elevated risk for hepatitis A (including PEH and PWUD) are not static; new susceptible individuals become part of these risk groups despite even adult vaccination efforts. Together, these patterns highlight persistent gaps in adult hepatitis A vaccination coverage that maintain population susceptibility and may increase vulnerability to future outbreaks.

We also examined where individuals were receiving hepatitis A vaccinations, since certain provider types and settings (particularly community health centers) are more accessible to at-risk populations and have historically been central to outbreak response efforts. These settings administered most adult hepatitis A vaccinations during the 2018-2020 outbreak period but did not return to pre-2020 vaccination volumes following the COVID-19 pandemic (**Fig 1B**). This sustained reduction suggests decreased capacity within key access points for delivering hepatitis A vaccines during future outbreaks.

### HAV outbreaks in 2018-2020 and 2023-2024 were unconnected

The similar demographic profiles observed across the 2018-2020 and 2023-2024 outbreaks raised questions about the relationship between these two outbreaks. To investigate this, we generated 241 high-quality whole genome HAV sequences from cases in MA, excluding one sequence that could not be matched to case information (**Table S1**). Before performing phylogenetic analysis, we used epidemiological data to confirm that these sequences were representative of the outbreaks as a whole. Successfully sequenced MA cases were well distributed across the epidemic curve (**Fig 1A**) and reported distributions of gender, age, race/ethnicity, and vaccination status among sequenced outbreak-associated cases were similar to those observed for all outbreak-associated cases in both 2018-2020 and 2023-2024 (**Table 1**). The geographic distribution of sequenced outbreak-associated cases (based on city of residence) was also similar to that of all outbreak-associated cases in MA; in both groups, cases were more concentrated in densely populated areas such as Boston (**Fig 1C-D**, **Fig S2**). Sequenced cases from NH were likewise well-distributed across the epidemic curve (**Fig 1A**), although only outbreak-associated NH cases were sequenced in this study.

We then performed phylogenetic analysis to assign sequences to established HAV subgenotypes. We found that sequences generated in this study fell into three divergent HAV subgenotypes (**Fig 2A**, **Table S1**). The majority of successfully sequenced cases from 2018-2020 belonged to subgenotype IIIA (MA: 212 IIIA, 11 IA, 3 IB; NH: 50 IIIA, 12 IB), while successfully sequenced cases from MA in 2023-2024 belonged predominantly to subgenotype IB (9 IB, 3 IA, 3 IIIA). The 2018-2020 subgenotype IIIA sequences from both MA and NH formed a single, genetically similar cluster, consistent with rapid, localized transmission. The predominance of subgenotype IIIA among 2018-2020 MA and NH cases was notable in the context of the nationwide 2016-2020 HAV outbreak, which was largely driven by subgenotype IB (among 126 publicly available VP1-2B sequences from 2016-2020, 1.6% were IIIA, 76.2% IB, and 20.6% IA) ^5^. Although a small number of MA cases from 2023-2024 also belonged to subgenotype IIIA, they were genetically distinct from the 2018-2020 IIIA cluster and did not meet epidemiologic criteria for outbreak-associated cases (**Fig 1A**, **Fig S3**, **Fig S4**).

**Figure 2.**
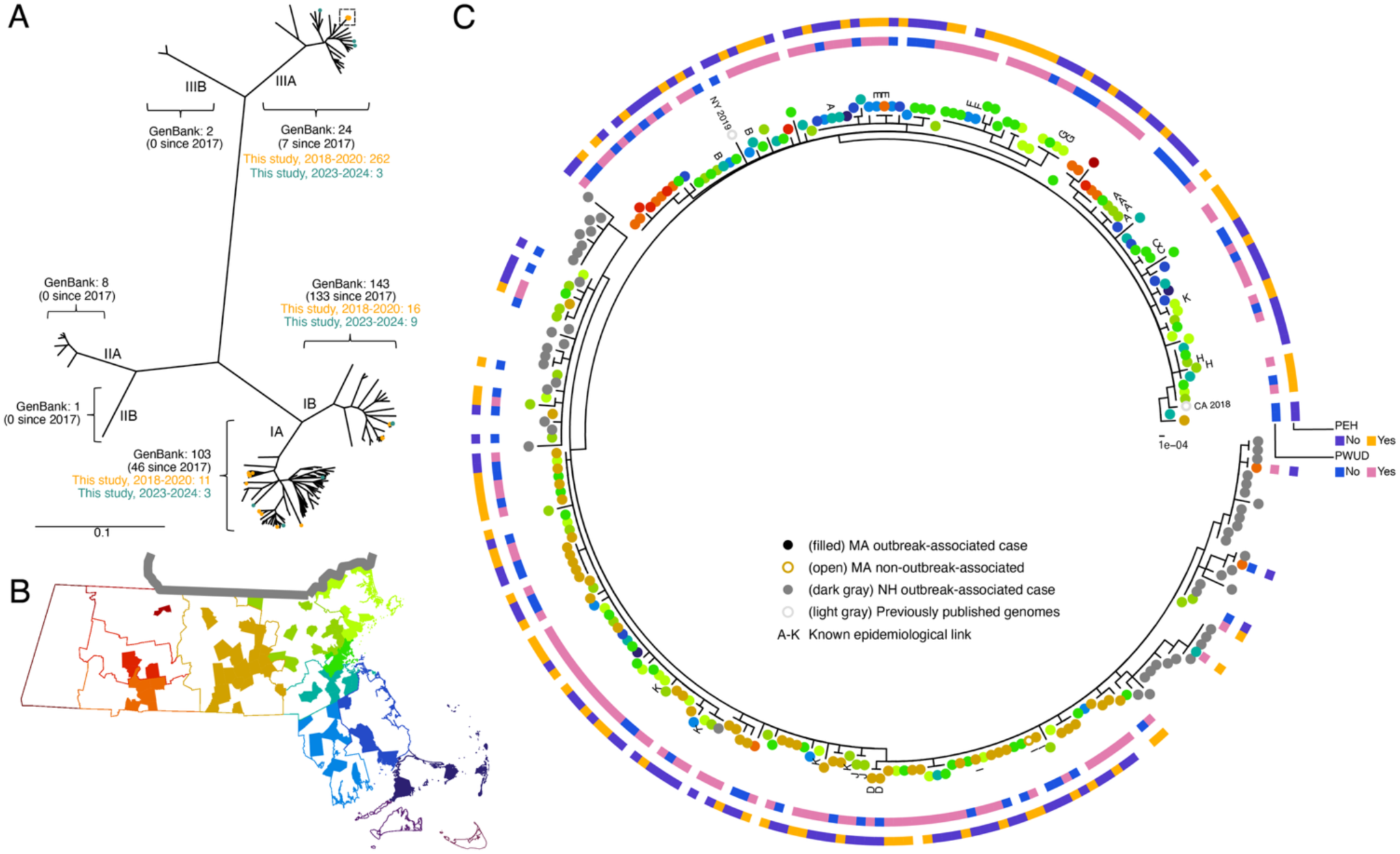
Clustering of 2018-2020 HAV subgenotype IIIA sequences by epidemiological and geographic variables. **A)** Unrooted phylogeny of all whole genome human host HAV sequences (see **Methods**), including both publicly available sequences and those sequenced in this study. Publicly available sequences with collection dates from 2017 or later are indicated. **B)** MA towns from which subgenotype IIIA cluster sequences generated in this study were derived, colored by county of residence (towns without sequence data are shown in white). Shapefiles used to create the map were downloaded from the Bureau of Geographic Information (MassGIS), Commonwealth of Massachusetts, Executive Office of Technology and Security Services database on 15 April 2024. The NH border is shown in gray above the MA state outline. **C**) Phylogeny of all sequences belonging to the 2018-2020 subgenotype IIIA cluster. MA counties are colored as in panel B, and all sequences from NH are shown in dark gray.

Likewise, subgenotype IA and IB sequences from the two outbreak periods did not form monophyletic groups on the phylogeny, further indicating that the 2018-2020 and 2023-2024 outbreaks were not directly linked.

Whole genome sequence data enabled closer examination of the genetically similar 2018-2020 subgenotype IIIA cluster and revealed patterns of intra- and inter-state spread. Although subgenotype IIIA cases were distributed across MA, individuals with identical HAV genomes were geographically closer to one another than individuals with non-identical genomes (**Fig S5A**), consistent with localized transmission (**Fig 2B-C**). Sequence data from NH cases allowed us to determine the extent of spread beyond MA and similarly showed localized transmission, though NH cases were interspersed among MA cases on the phylogeny, especially with cases from nearby northeastern MA (**Fig 2B-C**). Travel history information collected by NH DPH further supported cross-border spread: among the 50 NH sequences in the subgenotype IIIA cluster, nine of ten individuals reporting recent travel outside NH had traveled to MA. In addition, most of the earliest hepatitis A cases in NH resided near the NH-MA border which, coupled with the later start of the outbreak in NH compared to MA, raises the possibility that the NH HAV IIIA-associated outbreak may have been seeded from MA. Two previously published sequences also clustered within this subgenotype IIIA group; one from California (CA) from 2018 (MN062167.1) and one from New York (NY) from 2019 (ON524418.1). The close relatedness between cases from MA, NH, NY, and CA suggests that there may have been cases in other US states that were also connected to this subgenotype IIIA cluster, for which whole genome sequencing data were not available.

Detailed epidemiological information collected by MA DPH allowed us to assess whether closely related (both genetically and geographically) cases had a plausible transmission link. We identified 44 epidemiologically confirmed pairwise links among MA cases, including clusters associated with a restaurant and a jail. These known epidemiologic links were supported by genomic data, with cases known to be linked clustering together on the phylogeny (**Fig 2C**). We also found some evidence for clustering of sequences associated with PEH (delta = 1.039, p-value = 0.01) or PWUD (delta = 0.626, p-value = 0.02) using the entropy-based Delta statistic ^20^ with a permutation test to determine significance (see **Methods**). These results suggest that transmission was more likely to occur within risk groups than between individuals with and without known risk factors. It does not, however, preclude transmission between groups, and indeed we see individuals both with and without risk factors within clusters. We also leveraged multiple geographic variables collected in MA to evaluate whether variables other than a patient’s reported city of residence—which may be unreliable for PEH—could better inform genomics-based analyses. We found that pairwise geographic distances, calculated from either the patient’s city of residence or from the city where a provider ordered a diagnostic test, differ significantly between genomically identical isolates (0 SNPs) compared to more diverse isolates (>0 SNPs) (**Fig S5**). This suggests that the city where a provider ordered a diagnostic test may serve as a useful proxy for where individuals were exposed to and transmitted HAV.

### Known risk factors drive HAV transmission

Having observed that epidemiological links clustered on the phylogeny, we applied additional genomic analyses to investigate factors associated with increased HAV transmission. Specifically, we used the transmission reconstruction tool JUNIPER ^21^, which integrates deep sequencing and temporal data to infer likely transmission links. Focusing on the subgenotype IIIA cluster, we then contextualized the transmission reconstruction results by overlaying links identified through epidemiological investigations. These epidemiological investigations identified 16 linked pairs among the 211 MA subgenotype IIIA cases that were successfully sequenced. These 16 are a subset of the 44 total pairs identified in MA cases (only these 16 pairs had both cases successfully sequenced), highlighting the value of contact tracing especially when sequencing is not performed or is not successful. Indeed, because only ∼40% of known outbreak-associated cases were sequenced (and likely only half of outbreak cases were ever reported ^8^) the likelihood of capturing *direct* transmissions using transmission reconstruction from genomic data was low; nevertheless, transmission reconstruction from genomic data provided support for direct transmission for six of these epidemiologically linked pairs. Additionally, in two instances the reconstruction inferred an alternative transmission pathway between two epidemiologically linked cases that involved a third sequenced case not linked through contact tracing, highlighting the complementary information provided by the two approaches (**Fig 3A**). We also confirmed that links supported by both epidemiologic investigation and transmission reconstruction exhibited a similar distribution of genetic distances, which was narrower than that observed among all pairs consistent with potential transmission (defined as contemporaneous case pairs within the same county) (**Fig 3B**). This finding is consistent with identified transmission pairs being members of the same transmission chain; however, we note it cannot conclusively demonstrate direct transmission between the identified individuals or rule out transmission among other individuals in the dataset.

**Figure 3.**
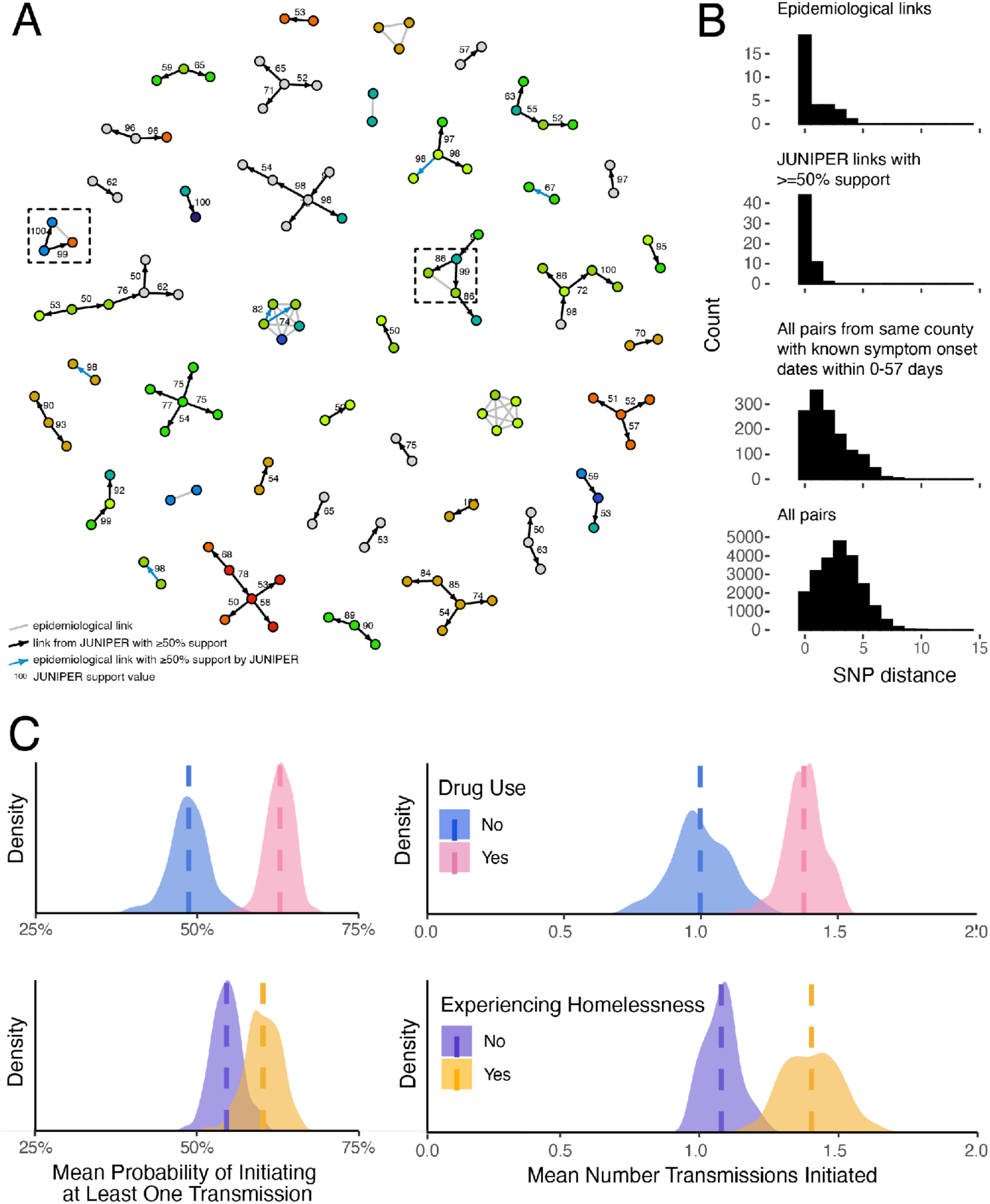
Transmission reconstruction of subgenotype IIIA cases using epidemiological investigation and genomic data. **A)** Transmission links identified through epidemiological investigation (MA only) or inferred by JUNIPER with ≥50% support (MA and NH) among successfully sequenced cases. Boxes highlight instances in which transmission reconstruction suggests an alternative infection pathway involving a third sequenced case not identified by epidemiological linkage. Node colors indicate MA county of residence, as in **Fig 2B**. **B)** For MA cases only, distributions of single-nucleotide polymorphisms (SNPs) for defined sets of case pairs. SNP distances between 0 and 15 are shown (see **Fig S6** for full distribution). **C)** For MA cases only, mean probability of initiating at least one transmission (left) and mean number of transmissions initiated across JUNIPER iterations (right) among PWUD/non-PWUD (top) and PEH/non-PEH (bottom). Density curves sum to 1.

The transmission links inferred by transmission reconstruction from genomic data allowed us to move beyond phylogenetic clustering by demographic characteristics and to examine the specific relationship between individual risk factors and transmission. First, we noted that individual transmission pairs identified from the genomic data included cases with and without PEH and PWUD risk factors, suggesting transmission not only within, but also between, risk factor groups. Next, we examined the importance of these risk factors to transmission. We evaluated the number of transmission events initiated by each 2018-2020 case in the subgenotype IIIA dataset across the distribution of transmission networks generated by JUNIPER and found that PWUD were associated with a higher mean number of transmissions initiated (1.38 in PWUD versus 1.00 in non-PWUD; p-value = <0.001). Individuals with any history of drug use were also associated with a higher probability of initiating at least one transmission event (63% in PWUD versus 49% in non-PWUD; p-value = <0.001) (**Fig 3C**). We observed a similar pattern among PEH, who showed a higher mean number of transmissions initiated compared with non-PEH cases (1.40 in PEH versus 1.08 in non-PEH; p-value = 0.011) and non-significant higher probability of initiating at least one transmission event (60% in PEH versus 55% in non-PEH; p-value = 0.065). By contrast, we observed a non-significant trend of decreased transmission from food handlers (**Fig S7**). To explore the potential impact of sampling biases, we repeated the transmission reconstruction under a range of sampling rates and found consistent results (**Table S2**). Ultimately, we conducted all transmission reconstruction analyses with a sampling rate of 40%, consistent with the proportion of outbreak-associated cases we sequenced.

### Tools and methods for future HAV surveillance

As part of this study, we assessed the utility of whole genome sequencing by comparing it to existing HAV subgenotyping approaches. Specifically, we compared a phylogeny constructed using full HAV genomes from all sequences in the subgenotype IIIA main cluster (**Fig 2C**) with a phylogenetic reconstruction generated using only the VP1-2B region from the same samples. We found that the whole genome phylogeny provided substantially greater resolution than the VP1-2B-based tree, which consisted of largely identical sequences (**Fig S8**). The impact of this loss of resolution was most evident when examining subgenotype IIIA MA genomes from individuals with known epidemiological links identified through MA DPH: while whole genome sequencing grouped together sequences from epidemiologically linked individuals, the analysis restricted to the VP1-2B region failed to resolve these links (**Fig 4A**), limiting our ability to reconstruct transmission networks and assess factors associated with HAV transmission.

**Figure 4.**
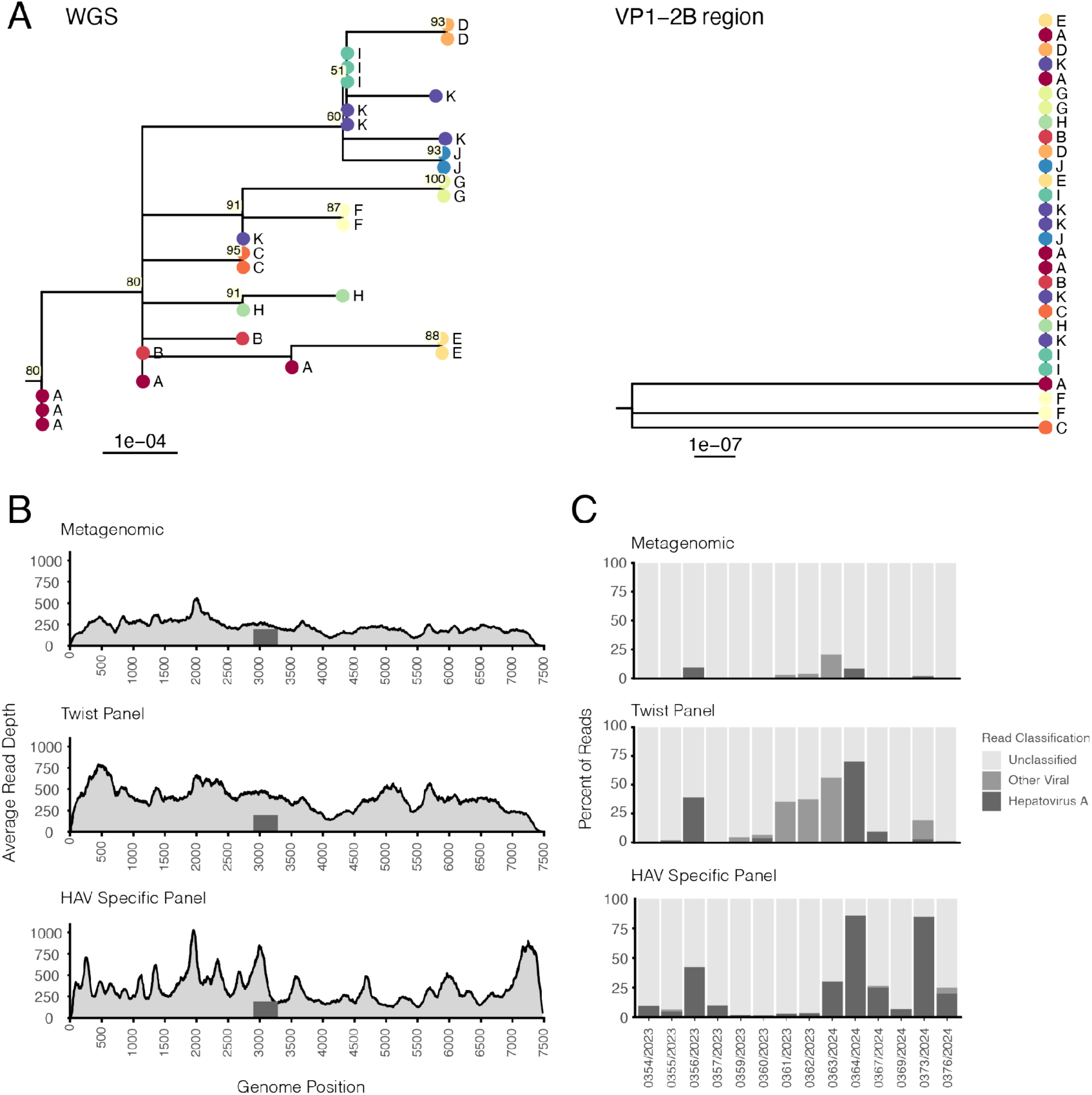
Comparison of HAV whole genome sequencing and subgenotyping approaches. **A)** Phylogenetic relationships among high-quality subgenotype IIIA genomes from this study with known epidemiological links reconstructed using whole genome sequences (left) and the VP1-2B region only (right). Linkage group designation is indicated by letters (A-K) and like colors, and numbers next to nodes indicate ultrafast bootstrap (UFBoot) values. **B)** Average sequencing coverage for samples (n=7) from the 2023-2024 outbreak period that produced full genome assemblies using each sequencing method. The VP1-2B region is marked by a dark gray bar. **C)** Percentage of reads per sample identified as HAV for each sequencing method.

We also evaluated whether untargeted whole genome sequencing approaches could be used to glean additional information, such as detection of coinfections, that may help contextualize HAV infection and transmission. To do this, we compared three laboratory approaches for HAV whole genome sequencing: fully unbiased metagenomic sequencing, multi-pathogen hybrid capture, and single-pathogen hybrid capture ^22^ (see **Methods**). We applied these methods to outbreak-associated samples collected by the MA DPH during the 2023-2024 outbreak period and found that all three methods successfully generated complete HAV whole genome sequences as defined in **Methods**, including full coverage of the VP1-2B region (**Fig 4B**, **Table S1**). As expected, hybrid capture-based methods resulted in a higher proportion of HAV reads per sample than metagenomic sequencing, with the HAV-specific capture panel outperforming the multi-pathogen panel on this metric (**Fig 4C**). While these differences have implications for sequencing efficiency and cost, the results demonstrate that HAV whole genome sequencing can be successfully performed using multiple laboratory strategies depending on surveillance goals and available resources.

Although the HAV-specific hybrid capture panel was most effective at enriching HAV reads, it is not a suitable method for identifying potential coinfections, which may be relevant to understanding an outbreak and to outbreak response. Coinfections were detected using both unbiased metagenomics and the multi pathogen hybrid capture panel. These included hepatitis B virus (HBV) (n=3), hepatitis C virus (HCV) (n=48), adeno-associated virus 2 (n=5), and GB virus C (n=108). Possible HIV reads were removed from raw data prior to analysis (see **Methods**) to comply with ethical approvals, though future investigation of HIV coinfection may be informative given the overlap in HAV and HIV risk groups. We confirmed 28 of 33 HCV and 1 of 1 HBV coinfections with available surveillance data, but we did not detect HBV and HCV in a number of patients with positive clinical tests for these viruses (**Table S3**), suggesting that limit of detection should considered when interpreting sequencing results.

Despite the utility of whole genome sequencing for identifying coinfections and revealing detailed transmission patterns, sequencing of the VP1-2B region remains a useful tool for high-level characterization of HAV outbreaks, particularly given the extensive public reference data available for this region. As of 3 February 2025, the National Center for Biotechnology Information (NCBI) GenBank ^23^ database included 1,773 VP1-2B sequences, compared to 281 whole HAV genomes. To facilitate genotyping and phylogenetic analysis using either whole genome or VP1-2B region data, we developed two publicly available tools for HAV analysis. First, we created Hepatypist, a stand-alone command line tool that efficiently assigns short HAV sequences to known subgenotypes and can be readily adapted for use with other hepatitis viruses ^24^. Second, we leveraged the Nextclade ^25^ platform to enable real-time visualization and subgenotyping of either whole genome or VP1-2B region sequences through a web-based interface, without requiring command line expertise. We compared results across both tools using data from CDC GHOST and previously developed ^26^ (but no longer maintained) HAV typing tools and found that both of our tools accurately identified HAV subgenotypes (**Table S4**). We hope these tools will facilitate future HAV outbreak investigations, including in settings with limited computational or bioinformatics capacity.

## Discussion

In this study, we integrated epidemiological and genomic data to distinguish and characterize hepatitis A outbreaks in MA and NH between 2018-2024. Epidemiological investigations revealed the community-driven nature of HAV outbreaks in 2018-2020 and 2023-2024, with a substantial portion of cases occurring in PEH and PWUD. Furthermore, analysis of statewide vaccination data revealed sustained declines in adult hepatitis A vaccination following the COVID-19 pandemic, suggesting ongoing population-level susceptibility that may increase vulnerability to future HAV outbreaks. Using genomic data, we demonstrated that cases identified during the 2023-2024 resurgence were unrelated to those from the 2018-2020 outbreak in MA, despite affected individuals sharing similar demographic characteristics and reported risk factors. Transmission reconstruction analyses indicated that PEH and PWUD were associated with increased HAV transmission, though genomic evidence supported HAV transmission between individuals with and without these risk factors. Using sequencing data from NH and publicly available genomes, we also showed that transmission likely extended across state borders. Because the vast majority of HAV cases are not sequenced at baseline, however, we were not able to draw inferences about the sources of the two outbreaks.

To support improved preparedness for future hepatitis A outbreak investigations, we also describe here multiple laboratory sequencing methods and computational tools that can be used to generate and analyze new HAV genomic data across a range of public health settings. Our typing tools, Hepatypist and the HAV Nextclade datasets, both enable subgenotyping from either whole genome sequences or VP1-2B-only data, though whole genome sequencing provided substantially greater within-outbreak resolution than analyses based on the VP1-2B region currently used for routine subgenotyping. Whole genome data are also necessary for identifying novel subgenotypes as they emerge, as existing subgenotyping approaches such as the CDC GHOST system rely on previously characterized VP1-2B signatures. Importantly, all three laboratory sequencing methods evaluated in this study were capable of generating complete HAV whole genome sequences, although pathogen-specific hybrid capture methods produced higher-quality assemblies than multi-pathogen or fully unbiased methods, as expected. However, pathogen-specific capture methods do not allow detection of viral coinfections, which may be relevant in populations at increased risk for multiple infections.

Characterizing the high rate of hepatitis C coinfection, for example, helped explain the unusually high morbidity and mortality associated with these outbreaks and provided useful information for targeting interventions (e.g., vaccine clinics could be co-located with other services for people with hepatitis C infection). While multi-pathogen methods and unbiased sequencing approaches offer this broader detection capability, further optimization is needed, as these methods did not consistently detect all clinically identified coinfections in our samples. Additional research is also needed to understand when during the course of infection different diagnostic methods are able to detect and characterize HAV and other infections.

The increased resolution provided by whole genome sequencing complemented traditional epidemiologic approaches and enabled us to explore drivers of HAV transmission in two different ways: by examining phylogenetic clustering by risk factors, and by reconstructing transmission networks. While transmission reconstruction allowed us to assess associations between specific risk factors and transmission, current reconstruction methods have important limitations ^21,27,28^. In particular, the tool used in this study relies on assumptions about transmission bottleneck sizes and evolutionary models that may not fully reflect HAV biology, and all reconstruction approaches are constrained by sampling biases and assumptions about unsampled individuals. Integrating transmission reconstruction with epidemiologic data, as done here, can mitigate some of these limitations, as epidemiologic investigations and contact tracing leverage social networks and contextual information and are not restricted to inferring direct transmission events. Specifically, we note that linked cases in our dataset included pairs with known direct epidemiological links and cases belonging to the same epidemiological cluster (see **Methods**), the latter of which may be especially susceptible to self-reporting inaccuracies and assumptions about case proximity. This likely explains discrepancies in **Figure 4A** (particularly the epidemiological linkage group depicted by the letter K, which does not cluster tightly on the phylogenetic tree), and highlights the value of combining epidemiological and genomic information to understand transmission patterns.

Both epidemiological investigation and transmission reconstruction identified experiencing homelessness and drug use as associated with increased HAV infections. However, determining whether these risk factors are drivers of transmission remains challenging: information such as drug use history is self-reported and may be incomplete, and available epidemiological information on this variable does not distinguish between current, active drug use and prior history of drug use— scenarios that may convey significantly different transmission risk. Additionally, case investigations may not capture all potentially relevant variables (e.g., variables we do not yet know may play a role in HAV transmission) or unreported cases ^8^. Nevertheless, our findings indicate that we should not consider at-risk groups as isolated, as HAV transmission occurred between individuals with and without reported risk factors and infections were not confined to specific populations. This has important public health implications, particularly as rates of homelessness have increased throughout the US within an adult population (those born before the hepatitis A vaccine was recommended as a routine childhood vaccination in 2006) that remains largely immunologically naive to HAV.

Identifying individuals who are infected with and transmitting HAV is essential for effectively and efficiently targeting public health interventions. In MA and NH, outbreak response efforts have primarily focused on targeted vaccination, though they have also included education, improving hygiene, and other outreach efforts. Determining where to deploy vaccines can be challenging when many affected individuals report unstable housing, as reported residential addresses may not accurately reflect where exposure or transmission occurred. Our analysis shows that the city in which a diagnostic test was ordered may be as informative as an individual’s reported city of residence when genetic similarity is used as a proxy for transmission. This suggests that ordering provider location may be a useful indicator of where vaccination efforts can be most effectively concentrated during hepatitis A outbreaks involving individuals with unstable housing, and this new finding should have broad utility for health departments investigating similar outbreaks nationwide. Because PEH may also have reduced access to insurance, nearby community health centers that provide vaccinations at no cost are likely to continue to play an important role in HAV outbreak response and should continue to stock hepatitis A vaccines, despite recent declines in vaccination delivery in these settings. Finally, because populations at increased risk for hepatitis A are dynamic—that is, new people join the group whenever drug use is initiated or housing becomes unstable—maintaining capacity for adult vaccination remains important even as cohorts vaccinated in childhood age into adulthood. Our new finding that PWUD and PEH are not simply more vulnerable to HAV infection, but may also drive transmission, underscores the importance of continued vaccination in these groups in order to prevent transmission and outbreaks in the broader population.

Expanding access to hygiene infrastructure for people experiencing homelessness has also been shown to play a role in controlling hepatitis A outbreaks elsewhere in the US ^29^. Accordingly, a combination of targeted vaccination and non-pharmaceutical interventions—such as increased access to public restrooms and showers, hand-washing stations, and hygiene kits—may help mitigate future hepatitis A outbreaks. Although these interventions, particularly the installation and maintenance of public restroom facilities, can be costly ^30^, the economic burden associated with HAV infection is also substantial ^31–33^, as is the burden of other infectious diseases that can be similarly mitigated with the same infrastructure investments. Additionally, monitoring of hepatitis A concentrations in wastewater is used by other states to enhance surveillance efforts and as an early indicator for future case increases, and could be a useful additional tool in monitoring community-wide hepatitis A ^9^ (of note, retrospective analysis of hepatitis A concentrations in Massachusetts wastewater shows an increased signal prior to the start of the 2023-2024 outbreak ^34^). These considerations underscore the value of using a combination of molecular, genomic, and epidemiological data to guide when and where interventions are deployed. We hope that our work demonstrating the utility of HAV whole genome sequencing for understanding outbreak spread and introducing accessible, open-source tools for HAV analysis will support more effective outbreak response and reduce the burden of hepatitis A, especially in our most vulnerable communities.

## Methods

### Ethical approvals

Sequencing of residual HAV samples from positive cases and analysis alongside associated epidemiologic metadata was reviewed by the Massachusetts Department of Public Health Institutional Review Board (IRB) (MA DPH IRB 00000701, #1377065 and #2143117) and covered by a reliance agreement at the Broad Institute.

### Sample collection and epidemiological investigation

In MA, all positive hepatitis A IgM results are reported electronically to the Massachusetts Virtual Epidemiologic Network (MAVEN ^35^), the state’s infectious disease surveillance and case management database. Reported cases are investigated via case and provider interview by local boards of health with support from DPH epidemiologists. Starting during the 2018-2020 outbreak, in addition to collecting clinical, demographic, and risk data, DPH epidemiologists conducting case investigations requested leftover serum specimens from the commercial and clinical labs that had performed the diagnostic testing, with a focus on cases that appeared to have been exposed within the US and were not close contacts of previously identified cases. Samples were sent to the Broad Institute, the MA State Public Health Laboratory (MA SPHL), and CDC’s Global Hepatitis Outbreak and Surveillance Technology (GHOST) for sequencing. One successfully sequenced sample was dropped from all analyses due to our inability to match the sample to epidemiologic data in MAVEN.

Epidemiological data for all confirmed and sequenced hepatitis A cases from 1 January 2018 through 31 December 2024 were extracted from MAVEN on 16 July 2025. Cases were classified using the Council of State and Territorial Epidemiologists (CSTE) Clinical Criteria of acute illness with a discrete onset of any sign or symptom consistent with acute viral hepatitis (e.g., fever, headache, malaise, anorexia, nausea, vomiting, diarrhea, abdominal pain, or dark urine) and either jaundice or elevated total bilirubin levels >3.0 mg/dL, OR, elevated serum alanine aminotransferase (ALT) levels >200 IU/L, AND, or the absence of a more likely diagnosis ^36^. To be considered “confirmed”, a case had to have met the CSTE clinical criteria and tested IgM anti-HAV positive, or had hepatitis A virus RNA detected (e.g., by PCR or genotyping). Alternatively, a case could have met the “confirmed” clinical criteria and occurred in a person who had contact (e.g., household or sexual) with a laboratory-confirmed hepatitis A case 15-50 days prior to onset of symptoms.

In the early stages of the 2018-2020 outbreak, cases were considered outbreak-associated only if they had known outbreak risk factors (i.e., unstable housing, drug use). However, in the context of large outbreaks nationwide, CDC issued guidance stating that a case should be considered outbreak-associated unless the person had traveled somewhere with endemic hepatitis A (and did not also report outbreak risk factors) or was part of a foodborne outbreak. This definition was applied retroactively as well as proactively to cover the full outbreak. In the 2023-2024 outbreak, the outbreak definition was narrower: an outbreak-associated case had known drug use, unstable housing, or incarceration.

Risk factors extracted from MAVEN for analyses included if a person was experiencing homelessness (PEH) at the time of infection and if the patient self-reported use of drugs in their lifetime (PWUD).

Drug use was defined as any use of illegal drugs not prescribed by a provider, including injection drugs and intranasal drugs, as well as drug use where method of use was non-injection, non-intranasal, or unknown. This definition was not meant to include cannabis use, although there may be some variability in how variables were explained and data collected. MSM were defined using sexual orientation and gender variables collected in MAVEN and reported sexual partners. Linked cases included pairs with known direct epidemiological links and all pairwise combinations of cases belonging to the same epidemiological cluster, as identified during case investigation.

Vaccination data were extracted from the Massachusetts Immunization Information System (MIIS), a confidential, web-based immunization registry that collects and stores the vaccination records of anyone who receives a vaccine in Massachusetts.

In NH, clinicians and laboratories reported suspected and confirmed HAV infections to the New Hampshire Division of Public Health (NH DPH) in accordance with state notifiable disease reporting requirements under RSA 141:C. NH DPH staff collected medical records, conducted patient interviews using hypothesis generating questionnaires, and identified risk factors for infection utilizing the New Hampshire Electronic Disease Surveillance System (NHEDSS). Similar to MA DPH, CSTE case definitions were applied to reported cases of HAV. Serum specimens were requested from all case-patients and sent to New Hampshire Public Health Laboratories (NH PHL) for testing.

### Nucleic acid extraction

Serum samples from MA patients who tested positive for HAV were extracted at MA SPHL using the MagNA Pure 96 DNA and Viral NA Small Volume Kit with 200µL of sample input and 50µL output.

Samples with sufficient volume from the 2023-2024 outbreak were extracted in duplicate for sequencing at both the Broad Institute and DPH. A water extraction was carried out with each extraction as a negative extraction control. Serum samples from NH patients who tested positive for HAV by PCR were extracted at NH PHL using the MagNA Pure Compact Kit.

### Illumina library preparation and sequencing

Sequencing for the 2018-2020 outbreak was carried out by the Broad Institute using a metagenomic sequencing approach. Briefly, these samples were treated with TURBO DNase using the TURBO DNA-free Kit (Thermo Fisher) and then RNase H to remove human ribosomal RNA as described previously ^37^. Samples underwent cDNA synthesis via a randomly primed reverse transcription and sequencing libraries were prepared with Illumina Nextera XT. External RNA Control Consortium (ERCC) RNA standard spike-ins were added to all samples sequenced at the Broad Institute to control for contamination.

Samples from MA from 2023-2024 (n=23 samples from 22 unique cases) were extracted in duplicate with one extraction sequenced by the Broad Institute and the second—when there was sufficient volume—by the MA SPHL. At the Broad Institute, samples were processed using a metagenomic approach as described above. After library preparation, samples were split so that the library was sequenced on the NextSeq 2000 platform with and without being subjected to the Twist hybrid capture comprehensive panel (**Table S1**). At MA SPHL, samples were treated with TURBO DNase using the TURBO DNA-free Kit (Thermo Fisher) with a DNase incubation time of 20 minutes and then sequenced using a previously designed HAV-specific hybrid capture panel ^22^. Hybrid capture libraries were prepared using the Illumina DNA Prep with Enrichment kit. Libraries were sequenced on the NextSeq 1000.

### Contextual genomes for assembly and analysis

Contextual genomes were retrieved from GenBank ^23^ on 5 March 2024 by searching for taxon identifier 12092 (Hepatovirus A, which includes all hepatitis A virus). Results were filtered to those with ≥7,125 unambiguous bases; human, sewage, or food hosts with known date and location; and no passage through cell lines or non-human animals. Collection date for sample MN062167.1 was retrieved through personal communication with authors. The latitude and longitude of provided cities, states, or countries were retrieved from gps-coordinates.net, with the most populous city used to represent states,countries, or continents.

### Hepatitis A genome assembly

HAV genomes were assembled on the Terra.bio platform using viral-ngs ^38^ as described below. The references used for assembly were selected from the contextual genomes above in order by sequence length to include only genomes with no ambiguous bases and ≥5% difference from already included genomes. The resulting sequences were: OK625565.1 (IA Haiti 2016), LC373510.1 (IA Japan 2014), KF773842.1 (IA Italy 2013), MK829707.1 (IB New Hampshire 2013), LC128713.1 (IB Thailand 2000), LC515201.1 (IB Gabon 2016), MH933768.1 (IIA Cameroon 2014), AY644670.1 (IIB Sierra Leone 1988), FJ360732.1 (IIIA India 2008), AB279733.1 (IIIA Japan 1992), AB258387.1 (IIIB Japan 1990). NC_001489.1 (IB), the RefSeq genome for Hepatovirus A, was also included.

Prior to assembly, human and HIV-1 reads were depleted using the deplete_only workflow in viral-ngs. HIV-1 reads removed were those that aligned to one of 350 representative HIV-1 genomes selected from among those included in the HIV-1 Nextclade dataset, accessed 2 December 2024. HIV-1 genomes were selected, in order from longest to shortest, if they had no ambiguous bases and were ≥5% different from already selected genomes. Additional parameters were as follows: bwaDbs=[“gs://pathogen-public-dbs/v0/hg19.bwa_idx.tar.zst”]; blastDbs=[“gs://pathogen-public-dbs/v0/GRCh37.68_ncRNA.fasta.zst", “gs://pathogen-public-dbs/v0/hybsel_probe_adapters.fasta”, “gs://pathogen-public-dbs/v0/metag_v3.ncRNA.mRNA.mitRNA.consensus.fasta.zst”, “gs://pathogen-public-dbs/v0/metagenomics_contaminants_v3.fasta.zst”].

Human- and HIV-1-depleted reads were then cleaned and contiguous sequences were assembled using the classify_single workflow in viral-ngs with no subsampling (specified by setting spades_n_reads to a large value, e.g. 1,000,000,000). Finally, HAV was assembled using the assemble_denovo workflow in viral-ngs with the cleaned reads produced by classify_single, the references described above, trim_clip_db (gs://pathogen-public-dbs/v0/contaminants.clip_db.fasta), and otherwise default parameters. Sequences with <7,125 unambiguous bases (<95% of 7,500 bases) or mean read depth <10x were excluded from downstream analyses.

To detect sub-consensus inter-sample contamination, Polyphonia ^39^ was run on all samples using the detect_cross_contamination_precalled_vcfs workflow in viral-ngs, with assemblies generated using the assemble_refbased workflow in viral-ngs with reference FJ360732.1 (lineage IIIA), intrahost variation detected using LoFreq through the isnvs_lofreq workflow in viral-ngs, read depths catalogued using the calc_bam_read_depths workflow in viral-ngs, and default parameters. Three samples were flagged as potentially contaminated (contamination volume ≥1%) and further excluded from downstream analyses: HepA/USA/MA/0052/2018, HepA/USA/MA/0070/2018, HepA/USA/MA/0238/2018. One additional sample, HepA/USA/MA/0142/2018, was also excluded from downstream analysis because its assembly did not match the consensus assembly generated from another sample from the same patient. In nine other pairs of samples taken from the same patient, all pairs were identical. In these cases, the sample with higher genome coverage and mean depth was included in subsequent analysis.

### Subgenotype assignment

The canonical GHOST typing region comprises the VP1-2B region of the HAV genome, i.e., positions 2,968 through 3,322 (1-indexed) relative to reference NC_001489.1. Subgenotypes were assigned to sequenced samples using this region using our newly developed Hepatypist tool ^24^ and Nextclade datasets ^40^.

To build the Nextclade datasets used for subgenotyping, all sequences with taxon ID 12092 were downloaded from NCBI GenBank ^23^ on 3 February 2025 and filtered to human hosts only via host field or, if that field was blank, using the isolation_source field and associated publication(s) in the title field. This set was then filtered by length to sequences with ≥7,125 unambiguous bases (≥95% HAV genome) to obtain the Nextclade whole genome sequencing (WGS) dataset (n=282), with the reference sequence set to accession NC_001489.1. We also created a VP1-2B Nextclade dataset for subgenotyping from the VP1-2B region only. To obtain this reference dataset, the human host-filtered taxon ID 12092 dataset was filtered to include only sequences overlapping the full VP1-2B typing region (n=1774). Of note, the WGS dataset does not include a representative sequence for subgenotype IC, as no IC WGS sequences existed in GenBank when the reference dataset was compiled.

Hepatypist is also packaged with its own VP1-2B reference dataset, which consists of deduplicated VP1-2B sequences extracted from the Nextclade WGS dataset plus added GenBank sequences representing subgenotype IC (total unique sequences = 142). Hepatypist subgenotyping relies on BLAST-based sequence metrics as well as phylogenetic placement using FastTree ^41^, as described in the Hepatypist wiki.

The Nextclade (WGS and VP1-2B) and Hepatypist datasets were assigned ground-truth subgenotypes using a separate Nextclade dataset constructed with unpublished VP1-2B reference sequences obtained from the CDC GHOST team, along with sequences for clades IIB (accession: AY644670.1) and IIIB (accession: AB279735.1).

### Maximum likelihood estimation and root-to-tip regression

All sequences passing quality control classified as IIIA from 2018-2020 were aligned using MAFFT v7.526 ^42^ along with the most recent complete (≥7,125 bases) IIIA genome from before 2018, AB973400.1 (collection date 31 January 2014) as an outgroup. A number of tree construction methods and substitution models were explored, as shown in **Table S5**. All trees were rooted using the outgroup sequence (AB973400.1) using the root function of the *ape* v5.8 package ^43^ in R v4.5.2.

The outgroup was then removed from the trees using the drop.tip() function in *ape* with resolve.root set to TRUE.

The best model and tree were selected by evaluating a root-to-tip regression and the distance between links identified through epidemiologic outbreak investigation. Within each linkage group identified through outbreak investigation, the mean pairwise tip distance within each linkage group was calculated for each tree using the cophenetic.phylo() function in the *ape* package in R. For each tree, the root mean square of the mean pairwise tip distances was calculated across linkage groups. IQ-TREE with GTR+I minimized the root mean square of the mean pairwise tip distances across linkage groups while maximizing the R^2^ of the linear best fit of the root-to-tip regression, and was therefore used for all analyses (though all trees generated by IQ-TREE were topologically similar). IQ-TREE –polytomy flag was used to collapse poorly resolved near-zero branches into multifurcated polytomies, and run with 1000 ultrafast bootstrap (UFBoot ^44^) replicates. Trees were visualized using the *ggtree* package in R ^45^.

The full unrooted phylogeny includes all contextual genomes described in the ‘Contextual genomes for assembly and analysis’ section above (103 IA, 143 IB, 8 IIA, 1 IIB, 24 IIIA, and 2 IIIB sequences) plus all new sequences from this study (266 IIIA, 25 IB, and 14 IA, minus one IIIA sequence that could not be matched to case information). Sequences were aligned using MAFFT v7.526 ^42^ and the phylogeny was constructed using IQ-TREE v2.3.4 ^46^.

The phylogeny in **Fig 2C** was constructed using all 2018-2020 IIIA sequences passing quality control (n=263), plus two publicly available IIIA sequences (MN062167.1 and ON524418.1) that clustered with them in the unrooted phylogeny described above and the IIIA outgroup used for rooting (AB973400.1). The outgroup and one sequence that we were not able to match with epidemiological case information were dropped after rooting as described above.

The phylogenies in **Fig 4A** were constructed using epidemiological links as defined in ‘Sample collection and epidemiological investigation’ above using AB279732.1 as an outgroup, which was dropped after rooting as described above. Several polytomy nodes were manually rotated for a clearer topological visual using the ggtree function flip().

### Time-resolved phylogenetic reconstruction

The time-resolved phylogeny was constructed using IQ-TREE v2.3.4 on a dataset including: all 2018-2020 IIIA sequences passing quality control minus the one sequence that could not be matched to case information (n=262) and the two publicly available IIIA sequences (MN062167.1 and ON524418.1) as described above. This was converted to a time tree using TreeTime v0.11.5 ^47^, with polytomy resolution via stochastic-resolve as recommended by the developers, and clock-filter adjusted to 0 to appropriately scale original input branch length. The two publicly available sequences were dropped for visualization using the *baltic* Python package ^48^ .

### SNP distances

Pairwise SNP differences were calculated as a raw count of unambiguous SNP calls. Only positions containing unambiguous bases were called as SNPs; all non-ATCG characters (gaps, Ns, etc.) were excluded from comparison and did not increment the distance count.

### Genome clustering by metadata variables

The entropy-based Delta statistic ^20^ was calculated to assess phylogenetic signal for PEH and PWUD using all 2018-2020 IIIA sequences passing quality control minus the one sequence that could not be matched to case information (n=262). All polytomies in the IIIA maximum likelihood phylogeny were resolved to bifurcations using the multi2di() function from *ape* ^43^. To force metric input, 0 branch lengths were disallowed by adding a small positive value (0.000000001) to each existing edge length. Tips with missing, “Unknown”, or “No” values for each variable of interest (PEH or PWUD) were encoded as “No/Unknown”. As a null distribution, the delta statistic was calculated ^20^ over 100 permutations of random shuffling of categorical metadata on the tree using a rate parameter of 0.1, proposal standard deviation of 0.05, and 50,000 iterations, keeping every 10th iteration, with a 500 iteration burn-in. The p-value of each test was calculated from a proxy: the probability of the random permutation delta being greater than the actual tree trait vector delta. The threshold for significance was set at p-value=0.05. Results were assessed for robustness across multiple prior distribution parameters.

### Transmission reconstruction

Transmission reconstruction was performed on clade IIIA genomes using the JUNIPER ^21^ software, which takes aligned consensus genomes, intrahost variants (iSNVs), and collection dates as input. iSNVs were identified and quantified using LoFreq within the viral-ngs assemble_refbased workflow. Consensus genomes used for transmission reconstruction (which requires all sequences to be assembled using a common reference) were assembled from raw reads using the assemble_refbased workflow in viral-ngs using the most commonly chosen IIIA reference from assemble_denovo (FJ360732.1) and otherwise default parameters. Read depths were calculated using the calc_bam_read_depths workflow in viral-ngs. Consensus genomes and reference sequences were aligned using MAFFT v7.511.

JUNIPER parameters were set as follows: generation interval (time between infections), 26.9 days ^49^, sojourn interval (time from infection to collection date), 34.2 days, calculated by adding the mean incubation period (28 days ^50^) and the mean time from symptom onset to sample collection among included samples with available symptom onset and collection dates (6.2 days). The mutation rate was fixed and set to 1.99 × 10^−4^ substitutions per site per year ^51^. JUNIPER was run five separate times, each with 50,000 global iterations: one with a varying, inferred sampling rate (92%) and four others with sampling rates fixed at 10%, 20%, 30%, and 40% (**Table S2**). The initial 20% of iterations were then discarded as burn-in and the resulting chain from each run was independently downsampled using the *coda* R package ^52^ to minimize autocorrelation between iterations. This resulted in effective sample sizes (ESSs) of 410, 100, 71, 104, and 92 respectively for each sampling rate.

Following this, direct transmissions were inferred using outputs directly from JUNIPER assuming a sampling rate of 40%. To calculate the expected number and probability of transmission between different metadata characteristics, we calculated the per-iteration expected number of descendants for each case under JUNIPER’s stochastic-epidemic model. The average number of descendants (or probability of any descendants), representing the number of transmissions (or probability of transmission), was then calculated for each iteration and the distributions across all iterations were then plotted. P-values were calculated as the percent of iterations in which the average number (or probability) of descendants in a given category was less than that in its comparator category (e.g., the number of iterations where PWUD transmitted to fewer people than non-PWUD, divided by the ESS).

### Detection of coinfections

Reads were taxonomically classified using Kraken2 v2.2.5 ^53^ through the classify_single workflow in viral-ngs. Viral species were catalogued that matched ≥10 reads in at least one sample, did not have “phage” in their names, and were listed as having human host in NCBI taxonomy browser or, if no host was listed, had a descendant with human host listed. Viral assembly was then run using reads that matched this set of viral species using the scaffold_and_refine_multitaxa workflow in viral-ngs with the cleaned reads and contiguous sequences produced by classify_single, RefSeq genomes as references, and otherwise default parameters. NCBI RefSeq genomes for each viral species were used as references, along with the diverse references previously described in ‘Hepatitis A genome assembly’ above. A species was determined to be detected in a sample if the resulting assembly included at least ≥1,000 bases. Assemblies of Zaire ebolavirus and RSV were determined to be lab contaminants from other sequencing activities and were excluded from the list of detected pathogens.

### Code availability

Code for all analyses and figure generation are available at: https://github.com/MASPHL-Bioinformatics/manuscript-HAVgenomics-2026 ^54^. There are no restrictions on access to code used for the analyses presented in this manuscript.

## Data Availability

Data approved by the IRB for public dissemination are available as Data S1. HAV sequence data generated as part of this study have been deposited in NCBI GenBank under BioProject accession PRJNA1442061 (BioSample accessions: SAMN56709630-SAMN56709934). Associated metadata are available in BioSample entries and in Table S1. In compliance with the IRB, only year of sample collection and outbreak association are included in line-level data; other relevant metadata variables are shared in aggregate in Table 1.

https://github.com/MASPHL-Bioinformatics/manuscript-HAVgenomics-2026

## Acknowledgements

We would like to thank all of the local health agents and MA and NH DPH epidemiologists who investigated individual hepatitis A cases described and sequenced here. We also would like to thank the GHOST team at the US Centers for Disease Control and Prevention for providing us with reference sequences, as well as Stephanie Ash and the MA SPHL Division of Virology for technical support and sample processing, and Elizabeth Daly, Zachary McCormic, Krystle Mallory, Kathrine Dzenis, Xinglu Zhang, and NH PHL Virology Staff. This publication was supported by the Office of Advanced Molecular Detection, Centers for Disease Control and Prevention through Cooperative Agreement Number CK22-2204 and by US National Institutes of Health (NIH) National Institute of Allergy and Infectious Diseases grant U19AI110818 (Genomic Centers for Infectious Diseases). Its contents are solely the responsibility of the authors and do not necessarily represent the official views of the Centers for Disease Control and Prevention.

## Author Contributions

L.A.K., L.B., B.L.M., G.R.G., S.S., D.J.P., and P.C.S. conceptualized the study. L.A.K., L.B., E. Decker, M.G., Z.T., and E. Dart curated data. L.A.K., L.B., E. Decker, M.G., Z.T., E. Dart, J.N., M.D., and S.W. conducted the formal analysis. L.C.M., P.C.S., and S.W. acquired funding. L.B., A.G-Y., K.S.M., A.O., J.L., C.L., K.C.D., K.J.S., D.C., C. Benton, K.H., H.B., C. Bean, E.A.T., and M.B. conducted investigations. L.A.K., L.B., T.M.B-F., M.G., Z.T., E. Dart, I.S., A.O., D.C., and M.B. contributed the methodology. K.C.D. and S.W. undertook project administration. F.G., S.B., N.E., G.R.G., S.S., E.F., and P.C.S. provided resources. L.A.K., T.M.B-F., M.G., and I.S. provided software. S.B., N.E., C.M.B., L.C.M., B.L.M., S.S., M.B., M.D., E.F., D.J.P., P.C.S., and S.W. supervised the study. L.A.K., L.B., M.G., Z.T., E. Dart, and J.N. performed visualization. L.A.K., L.B., T.M.B-F., E. Decker, M.G., Z.T., E. Dart, J.N., and S.W. wrote the original draft of the manuscript. All authors reviewed and edited the manuscript.

