## Supplementary Information for "Genomic epidemiology as a tool for understanding drivers of hepatitis A community outbreaks in Massachusetts and New Hampshire"

**Table S1.** Sequencing metrics for all HAV specimens sequenced in this study.

**Data S1.** Phylogenetic tree files and shapefiles required as inputs for analyses.

### Supplementary Figures and Tables

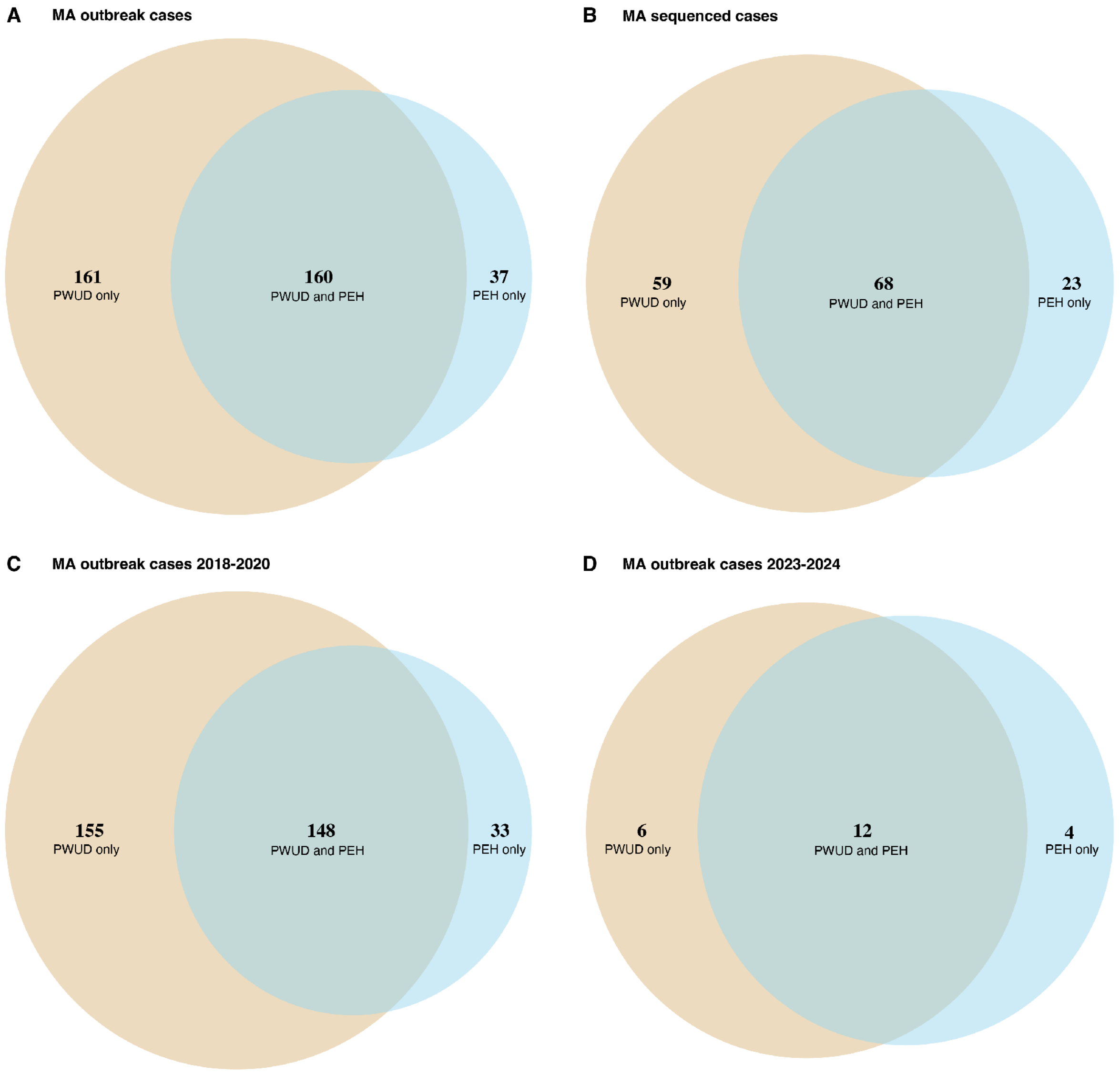


**Fig S1. Overlapping risk factors for hepatitis A.** Venn diagram of people who use drugs (PWUD) and people experiencing homelessness (PEH) for **A)** confirmed MA outbreak cases and **B)** all successfully sequenced MA cases of hepatitis A. MA outbreak cases by risk factor are further broken down into **C)** the 2018-2020 outbreak and **D)** the 2023-2024 outbreak.

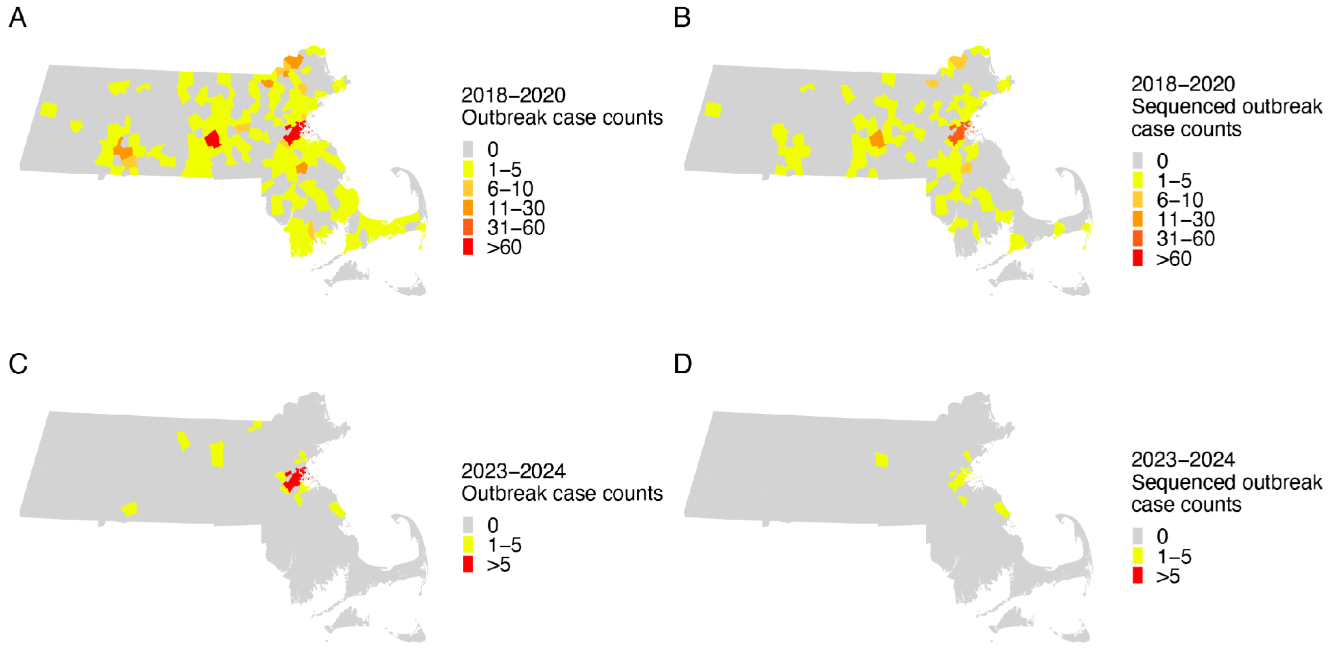


**Fig S2. Confirmed MA outbreak and sequenced outbreak case counts by outbreak period and town. A)** Confirmed outbreak-associated case count by MA town of residence during the 2018-2020 outbreak period. **B)** Sequenced outbreak-associated cases by town of residence for the 2018-2020 outbreak period. **C)** Confirmed outbreak-associated case count by town for the 2023-2024 outbreak period. **D)** Sequenced outbreak-associated cases by town for the 2023-2024 outbreak.

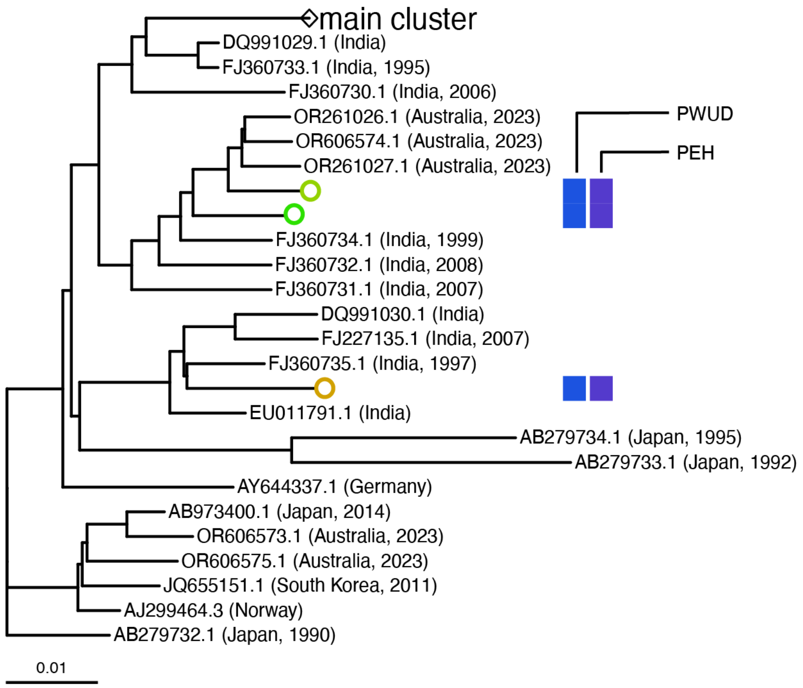


**Fig S3. Maximum likelihood phylogeny of clade IIIA sequences.** Phylogenetic reconstruction of all previously published (see **Methods**) and newly generated IIIA human host whole genome sequences passing quality control, with large genetically similar cluster shown in **Fig 2C** collapsed. Nodes from sequences generated in this study are colored by location and outbreak status as in **Fig 2C**; colored tips in this tree represent non-outbreak-associated sequences from 2023-2024. Colored bars to the right of the tree are also colored as in **Fig 2C**, indicating none of the newly generated MA sequences were from PWUD or PEH cases.

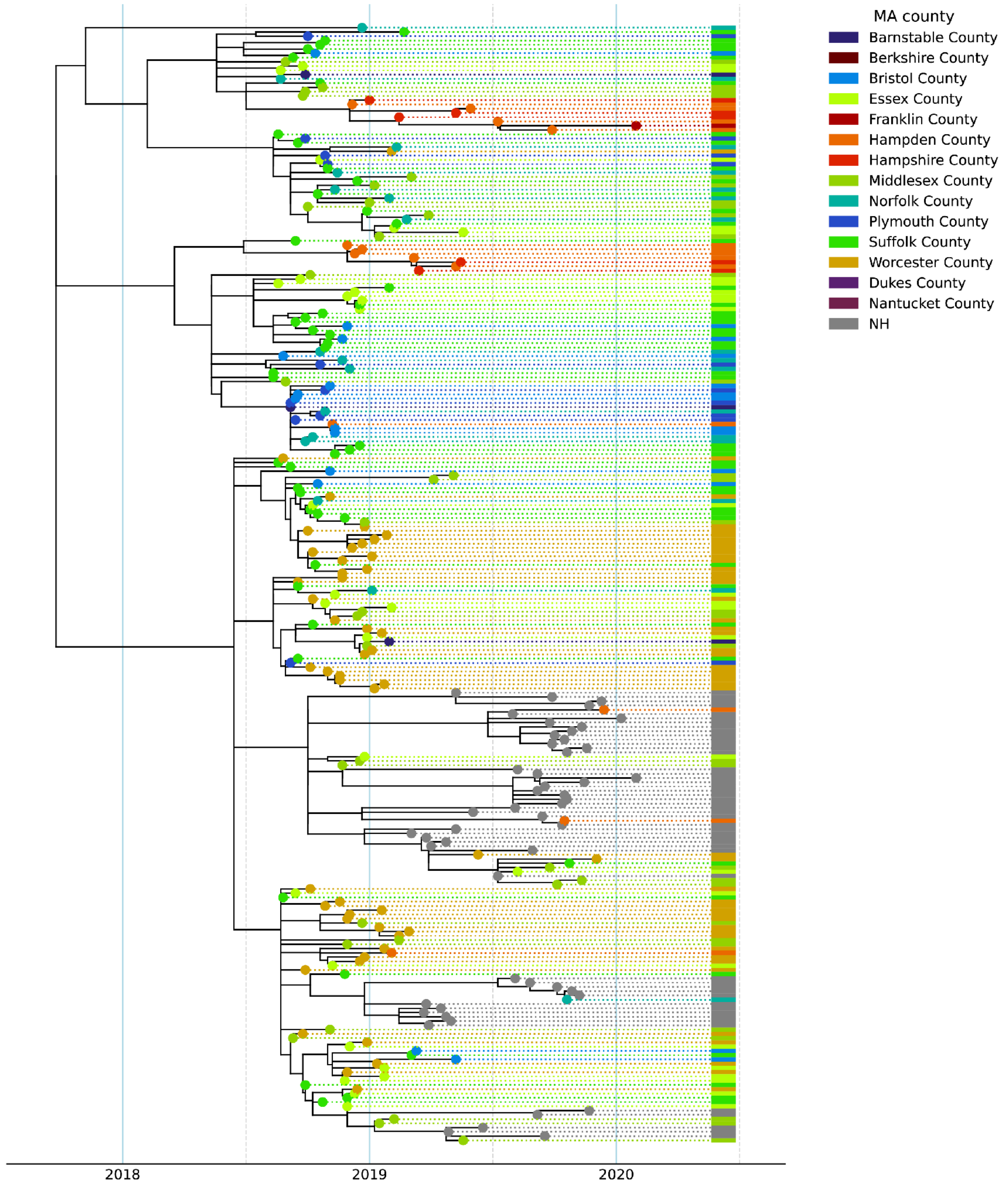


**Fig S4. Time-resolved phylogeny of subgenotype IIIA sequences.** IIIA sequences from the 2018-2020 outbreak in both MA and NH. Tips are colored by reported residence location (state or MA county).

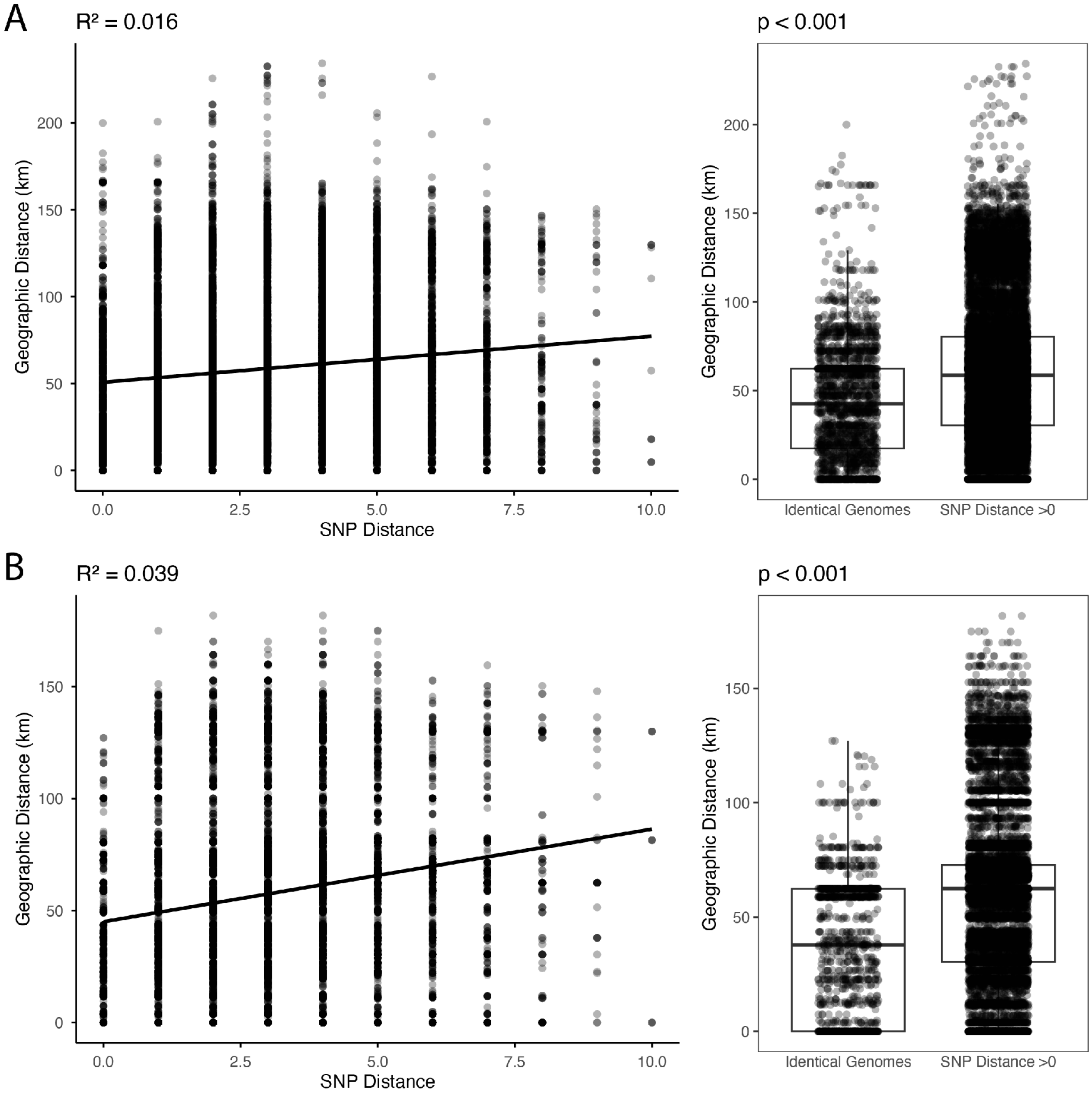


**Fig S5. Association between geographic distance and SNP distance for MA IIIA sequences.** Association between geographic distance and genomic distance, as measured by SNP distance, for MA IIIA cases passing quality control from 2018-2020. **A)** Geographic distance as measured between city of residence recorded for each patient sample. **B)** Geographic distance as measured between city where a provider ordered a diagnostic test for each patient sample.

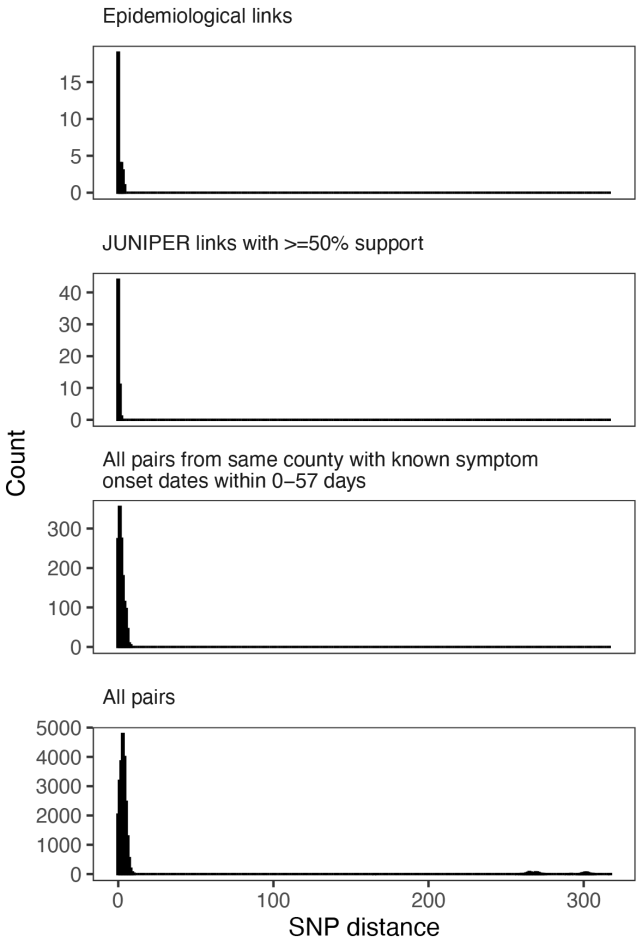


**Fig S6. SNP distances between pairs of MA IIIA sequences.** Full SNP distance distributions between pairs of sequences identified as **A)** epidemiological links, **B)** transmission events identified by JUNIPER with ≥50% support, **C)** all pairs of samples from the same MA county of residence with known symptom onset dates within 57 days, and **D)** all pairs of samples. Only IIIA MA sequences passing quality control are included.

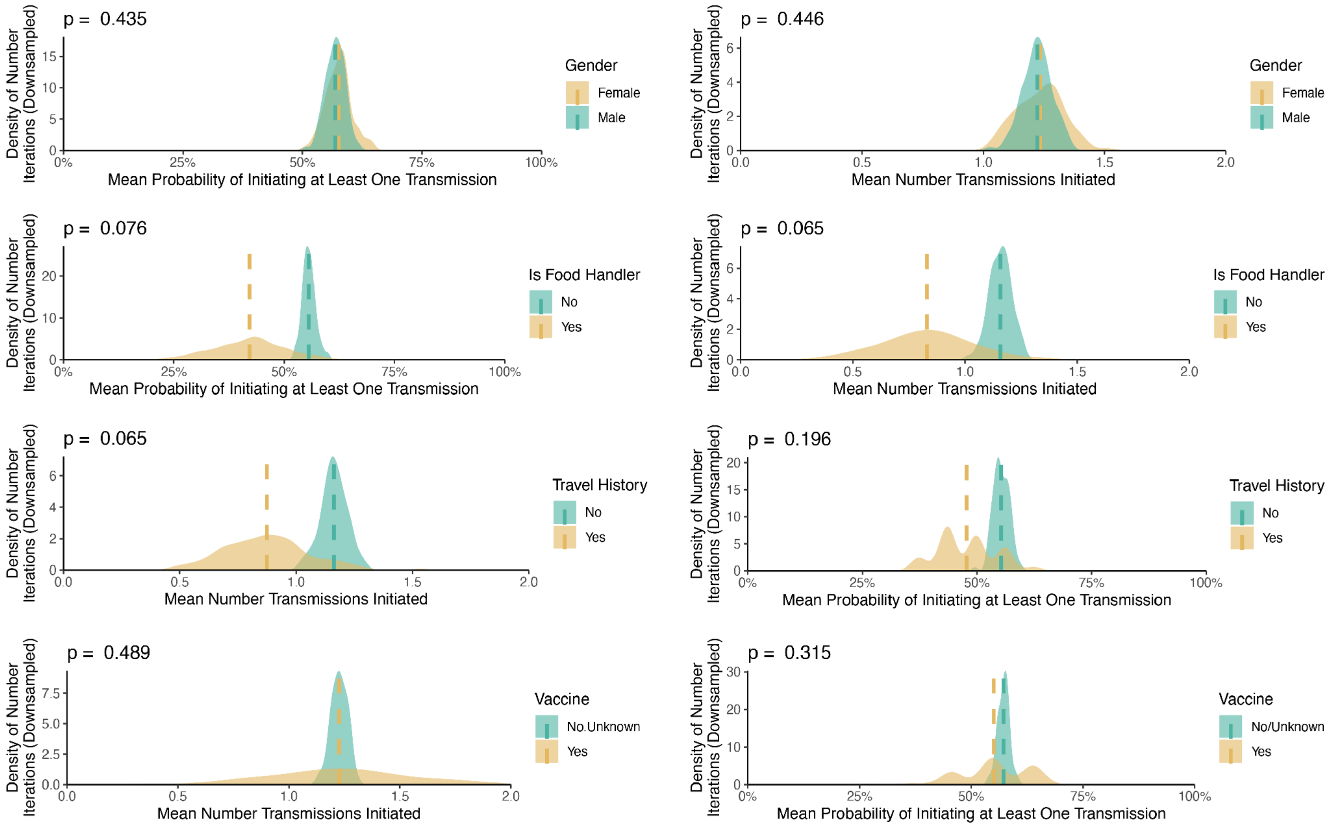


**Fig S7. Transmission probability by variable.** Transmission identified by reported gender, food handler status, evidence of travel history, and vaccination status. Only IIIA MA sequences passing quality control are included.

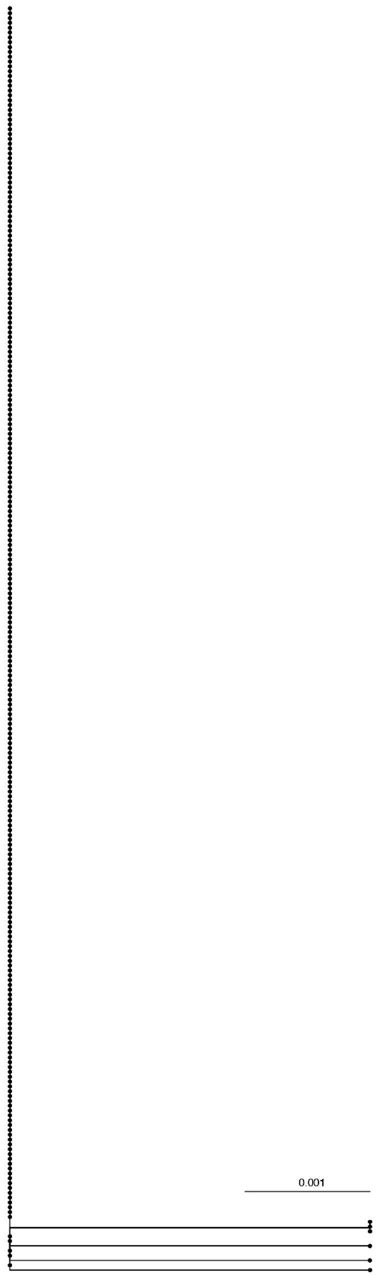


**Fig S8. Maximum likelihood phylogeny of all IIIA 2018-2020 VP1-2B region sequences.** Identical genomes are shown as dots on the vertical branch.

| From | To | Support Value for Transmission | | | | |
|---|---|---|---|---|---|---|
| | | Sampling Rate 40% | Sampling Rate 30% | Sampling Rate 20% | Sampling Rate 10% | Sampling Rate 92% |
| HepA/USA/MA/0030/2018 | HepA/USA/MA/0054/2018 | 67 | 58 | <50 | <50 | 89 |
| HepA/USA/MA/0055/2018 | HepA/USA/MA/0081/2018 | 98 | 98 | 96 | 93 | 100 |
| HepA/USA/MA/0122/2018 | HepA/USA/MA/0139/2018 | 98 | 92 | 96 | 85 | 100 |
| HepA/USA/MA/0224/2018 | HepA/USA/MA/0233/2018 | 74 | 54 | <50 | <50 | 100 |
| HepA/USA/MA/0224/2018 | HepA/USA/MA/0047/2018 | 82 | 79 | 76 | 62 | 100 |
| HepA/USA/MA/0278/2019 | HepA/USA/MA/0284/2019 | 98 | 95 | 94 | 85 | 100 |

**Table S2.** JUNIPER support values for transmission events identified by both epidemiological investigation and JUNIPER with ≥50% support using fixed sampling rates of 40% (used in all other JUNIPER analyses), 30%, 20%, or 10%, or the JUNIPER automatically detected sampling rate (92%).

| Coinfection | Genomic Detection | Diagnostic Test Detection | Count |
|---|---|---|---|
| Hepatitis C virus | Yes | Yes | 28 |
| | | No | 3 |
| | | Unknown | 2 |
| | No | Yes | 58* |
| | | No | 273 |
| | | Unknown | 11 |
| Hepatitis B virus | Yes | Yes | 1 |
| | | No | 0 |
| | | Unknown | 0 |
| | No | Yes | 8 |
| | | No | 365 |
| | | Unknown | 1 |

* Of 58 specimens for which the most recently reported HCV result included a positive PCR result, 32 were from the same week as the sequenced specimen.

**Table S3.** Hepatitis C virus and hepatitis B virus coinfections detected in MA HAV samples from genomic data and from PCR diagnostic tests (concordance = green; non-concordance = red), including in samples from which we were not able to generate a high-quality HAV whole genome sequence.

| | GHOST | | | GenBank | | |
|---|---|---|---|---|---|---|
| | **Concordant** | **Discordant** | **Unassigned** | **Concordant** | **Discordant** | **Unassigned** |
| **Hepatypist** | 553 | 0 | 0 | 93 | 0 | 0 |
| **HAVnet** | 551 | 1* | 1** | 92 | 0 | 1† |
| **Nextclade VP1-2B** | 553 | 0 | 0 | 93 | 0 | 0 |
| **Nextclade WGS** | 552 | 1* | 0 | 93 | 0 | 1† |

* One GHOST IC sequence was assigned IA by both HAVnet and Nextclade WGS. This was expected, as neither tool had an IC sequence in the reference set.

** One GHOST IA sequence was reported unassignable by HAVnet.

† The GenBank sequence KJ436954.1, clustering with IB on the GHOST reference tree, was reported unassignable by both HAVnet and Nextclade WGS. This test sequence is on the longest branch within the GHOST and Nextclade VP1-2B region clusters, which likely indicates additional site mutations or potentially recombination.

**Table S4.** Validation results for four tools run against two curated VP1-2B sequence sets. The first (“GHOST”) consists of reference sequences provided by the CDC GHOST team for testing. The second (“GenBank”) consists of 93 unique HAV sequences from GenBank that were not included in the Nextclade (WGS or VP1-2B) and Hepatypist reference datasets.

| Tree Software and Nucleotide Substitution Model | Root-to-Tip Association ($R^2$) | Linkage Group Mean Pairwise Tip Distance | $R^2$ / Linkage Group Distance Ratio |
|---|---|---|---|
| FastTree v2.1.11 [1] GTR + CAT + Gamma | 0.648 | 2.24e-04 | 2,897 |
| FastTree v2.1.11 [1] GTR + CAT | 0.648 | 2.10e-04 | 3,084 |
| FastTree v2.1.11 [1] JC + CAT | 0.629 | 1.83e-04 | 3,448 |
| IQ-TREE v2.3.4 [2] GTR + G4 | 0.460 | 1.59e-04 | 2,890 |
| IQ-TREE v2.3.4 [2] GTR | 0.424 | 1.72e-04 | 2,465 |
| IQ-TREE v2.3.4 [2] GTR + I + R | 0.390 | 2.10e-04 | 1,852 |
| IQ-TREE v2.3.4 [2] GTR + I | 0.484 | 1.38e-04 | **3,500** |
| IQ-TREE v2.3.4 [2] JC | 0.495 | 1.61e-04 | 3,074 |
| IQ-TREE v2.3.4 [2] HKY | 0.424 | 1.66e-04 | 2,550 |
| IQ-TREE v2.3.4 [2] ModelFinder [3] (TN + F + I) | 0.345 | 2.31e-04 | 1,493 |
| RAxML v4.0 [4] GTR + Gamma (1,000 bootstraps) | 0.475 | 1.76e-04 | 2,707 |
| MrBayes v2.2.4 [5] GTR + Gamma (1,100,00 iterations, 100,000 burn-in) | 0.139 | 1.18e-04 | 1,173 |

**Table S5.** Phylogenetic tree construction methods and substitution models explored. JC = Jukes-Cantor [6], TN = Tamura and Nei [7], HKY = Hasegawa-Kishino-Yano [8], GTR = general time-reversible [9], G = discrete gamma [10], I = invariant sites [11] , R = FreeRate generalization of gamma [12], CAT = categorical model [13].
